# Noninvasive Epileptogenic Signal Direction Determination by Effective Connectivity of Resting State Functional MRI - Independent of EEG

**DOI:** 10.1101/2022.01.31.22269916

**Authors:** Varina L. Boerwinkle, Bethany L. Sussman, Sarah N. Wyckoff, Iliana Manjón, Justin M. Fine, P. David Adelson

## Abstract

The goal of this study was to determine resting state fMRI effective connectivity (RSEC) capacity, agnostic of epileptogenic events, in distinguishing seizure onset zones (SOZ) from propagation zones (pZ). Consecutive patients (2.1-18.2 years old), with epilepsy and hypothalamic hamartoma, pre-operative rs-fMRI-directed surgery, post-operative imaging, and Engel class I outcomes were collected. Cross-spectral dynamic causal modelling (DCM) was used to estimate RSEC between the ablated rs-fMRI-SOZ to its region of highest connectivity outside the HH, defined as the propagation zone (pZ). RSEC between the SOZ and PZ was characterized as positive (excitatory), negative (inhibitory), or null. It was hypothesized that connectivity from the SOZ would be excitatory and connectivity from the pZ would be inhibitory. Sensitivity, accuracy, positive predictive value were determined for node-to-node connections. A Parametric Empirical Bayes (PEB) group analysis was performed to identify effects of Engel class outcome and age. RSEC strength was also evaluated for correlation with percent seizure frequency improvement, sex, and region of interest size. Of the SOZ’s RSEC, only 3.6% had no connection of significance to the pZ when patient models were individually reduced. Among remaining, 96% were in expected (excitatory signal found from SOZ→pZ and inhibitory signal found from pZ→SOZ) versus 3.6% reversed polarities. Both polarity signals were equivalently as expected, with one false signal direction out of 26 each (3.7% total). Sensitivity of 96%, accuracy of 93%, and positive predictive value of 96% in identifying and differentiating the SOZ and pZ. Groupwise PEB analysis confirmed SOZ→pZ EC was excitatory, and pZ→SOZ EC was inhibitory. Patients with better outcomes (Engel Ia vs. Ib) showed stronger inhibitory signal (pZ→SOZ). Age was negatively associated with absolute RSEC bidirectionally, but had no relationship with Directionality SOZ identification performance. This study demonstrates the accuracy of Directionality to identify the origin of excitatory and inhibitory signal between the surgically confirmed SOZ and the region of hypothesized propagation zone in children with DRE due to a HH. Thus, this method validation study in a homogenous DRE population may have potential in narrowing the SOZ-candidates for epileptogenicity in other DRE populations and utility in other neurological disorders.

**Highlights:**

- Directional rs-fMRI connectivity identifies seizure onset zone independent of EEG
- Signal from seizure onset zone to propagation zone is excitatory
- Signal from propagation zone to seizure onset zone is inhibitory
- Greater inhibition from propagation zone is associated with better surgical outcome

## 1. Introduction^1^

The most effective and only known curative treatment for drug resistant epilepsy (DRE) is surgery to remove the seizure focus and or interrupt the epileptogenic network (Luders et al., 2006). The primary determinant of surgical candidacy and success is accurate localization of the seizure onset zone (SOZ) (West et al., 2019). However, current noninvasive SOZ localization methods still often depend on confirmation by intracranial electroencephalograph (iEEG), which is expensive, carry risks, and still only leads to 40-80% seizure-freedom when a seizure focus is “identified” (Tonini et al., 2004).

Improvement in noninvasive SOZ source localization over more standard methods has been demonstrated by static connectivity from resting state fMRI (rs-fMRI) via independent component analysis (ICA) (Chakraborty et al., 2020). Notably, rs-fMRI ICA-derived SOZ findings have not only been associated with iEEG, but also with increased surgical candidacy. Additionally, improved Engel outcomes, have been associated with resolution of post-operative rs-fMRI SOZ ICA networks (Boerwinkle et al., 2019b; Boerwinkle et al., 2020; Boerwinkle et al., 2017). However, after evaluating over 2000 rs-fMRI of individual studies with DRE using ICA for SOZ localization, a major weakness remains: this *static* network measure identifies more than one plausible SOZ candidate in at least 40% of subjects (VB). Thus, there remains dependency on stereotactic iEEG to distinguish the best surgical target a concordance of data from rs-fMRI and other noninvasive methods.

The fundamental limitation of static rs-fMRI ICA is overcome to some extent by employing simultaneous EEG-fMRI, though this method only identifies the SOZ in a small proportion of cases, alone (Vaudano et al., 2021). The interictal epileptogenic discharges (IED) detected by EEG inform the time point at which pre-to-post blood oxygenation level dependent (BOLD) imaging maximal regions of interest (IED-BOLD ROI) are identified. A possible advance on this method, subsequent dynamic causal modeling (DCM) of the IED-BOLD ROI, has *differentiated the epileptogenic driver from propagation regions in four cases as determined by good surgical outcomes* (Engel class I and II) (Engel, 1993; Vaudano et al., 2021). However, this limited series highlighted the dependency of EEG-fMRI on requires additional specialized equipment, EEG technician staff, and epileptiform events to occur during the scan; all of which are cumbersome features limiting clinical utility.

To overcome these limitations, we designed a computational approach, Directionality, which employs cross-spectral DCM from *rs-fMRI alone, without electrophysiological information*, known as resting state effective connectivity (RSEC) helps to differentiate the driving SOZ region from propagation regions. These areas can be initially identified from the static rs-fMRI analysis. Directionality assumes SOZ are: First, (1) generators of excitatory (positive) signal toward regions of propagation; and secondly (2) receivers of inhibitory (negative) activity from the propagation node(s).

To determine if Directionality accurately distinguishes between SOZ and propagation zones (pZ), we selected a DRE population with homogenous and established SOZ localization, as well as prior surgically validated static rs-fMRI SOZ and pZ determined by the static rs-fMRI connectivity measure, SearchLight (SL) (Boerwinkle et al., 2018a). DRE from our patients with hypothalamic hamartoma (HH-DRE) fills these criteria because (1) it is well-established that the HH is the primary seizure driver in HH-DRE; (2) the HH epileptogenic network dynamic follow the Papez circuit, as verified by prior iEEG, rs-fMRI by ICA and partial correlation (Boerwinkle et al., 2016) and SL (Boerwinkle et al., 2018a), and EEG-fMRI by DCM (Murta et al., 2012; Usami et al., 2016). Notably, the same pathway of seizure progression from the HH to the rest of the brain from iEEG was found by rs-fMRI static measures alone in HH, but only on a group level (Boerwinkle et al., 2016) inferring the potential success of SOZ-identifying information from rs-fMRI Directionality on an individual basis.

In this study, we tested our hypotheses of signal direction by Directionality to distinguish the known HH’s SOZ from an ROI of propagation (the pZ) in children with HH-DRE who underwent pre-operative rs-fMRI SL-guided laser interstitial thermal ablation therapy (LITT) and had Engel I surgical outcomes. Further, we hypothesized that higher strength of signal from the identified region with negative signal (the pZ) and higher excitation from the region with positive signal (the SOZ) will correlate with seizure outcome. Also, because length of time of DRE is associated with decreased improvement with surgery (Tonini et al., 2004), we hypothesized that age will be associated with signal strength between these regions.

## 2. Methods

### 2.1 Participants

The local institutional review board granted approval for this study. Rs-fMRI became part of the standard preoperative evaluation for epilepsy surgery in 2012 and 2017 at Texas and Phoenix Children’s Hospitals, respectively, where the data was collected, therefore, no additional consent was deemed necessary for this retrospective rs-fMRI algorithm evaluation study.

Overall, there were 46 consecutive HH-DRE patients who underwent LITTs of pre-operative rs-fMRI surgical target, as in Boerwinkle et al. (2018a). Of these, 36 had Engel class 1 outcomes at least one year from surgery, meeting study criteria. Eight total patients’ data were excluded: four due to file corruption, one from partial data acquisition, and three from inadequate signal quality secondary to patient motion, yielding 28 total surgeries analyzed. Three patients, who had Engel 1b outcomes, subsequently had a second surgery, so both surgeries were included in the analyses. Rs-fMRI SOZ ablation location was confirmed by two blinded study personnel based on visualization of the pre-operative SOZ and post-operative imaging, as previously described (Boerwinkle et al., 2018a) (see **Supplementary Table 1**). The Directionality analyses were carried out by two rs-fMRI experts (BS and VB).

### 2.2 MRI Data acquisition and processing

#### 2.2.1 MRI acquisition

Images were acquired on a 3 Tesla MRI (Ingenuity, Philips Medical Systems, Best, Netherlands) equipped with a 32-channel head coil. Rs-fMRI parameters included TR (repetition time) 2000 ms, TE (echo time) 30 ms, matrix size 80×80, flip angle 80, number of slices 46, slice thickness 3.4 mm with no gap, in plane resolution 3×3 mm, inter-leaved acquisition, and number of total volumes 600, in two 10 minute runs, totaling 20 minutes. For anatomical reference, a T1-weighted turbo field echo whole-brain sequence was obtained with the following parameters: TR 9 ms, TE 4 ms, flip angle 8, slice thickness 0.9 mm, and in-plane resolution 0.9×0.9 mm.

#### 2.2.3 MRI preprocessing

Using the program Statistical Parametric Mapping version 12 (Friston et al., 1994) (SPM12; www.fil.ion.ucl.ac.uk/spm, Wellcome Trust Centre for Neuroimaging, London, UK), in Matlab 2019b, the T1 image was resampled to 1×1×1 mm voxels, the origin was set at the anterior commissure and, if needed, minor adjustments were made to correct orientation (head-tilt). The T1 image was segmented using the Computational Anatomy Toolbox version 12.7 (CAT12) (Gaser and Dahnke, 2016) and the Automatic Anatomical Labelling Atlas 3 (AAL3) (Rolls et al., 2020) was registered into subject T1 space and masks for each of the hippocampi were generated.

For the fMRI data, the same pre-processing steps as prior work (Boerwinkle et al., 2019b; Boerwinkle et al., 2017) were applied by high-pass filtering the data to remove ultra-low-frequency non-neural artifacts and extract the grey matter voxel time-course while removing voxels in the cerebrospinal fluid, and correcting for subject movement. Following ICA-based denoising (manual expert classification with removal of noise-based components in epilepsy (Boerwinkle et al., 2019b; Boerwinkle et al., 2017), CSF, and motion regression), the denoised functional data were re-aligned in SPM to register both of each subjects’ 10-minute rs-fMRI runs to each other. The mean functional image was segmented with unified segmentation. Using the bias-corrected mean functional image, the origin was set and orientation adjustments were made as described above for the T1. Functional images were co-registered (estimate) to T1 space in SPM using the bias-corrected mean functional image, with visual inspection.

#### 2.2.4 ROI Selection

Network ROI locations selected were: (1) the SOZ located by prior clinical SL and visibly ablated as shown in the immediate post-operative diffusion weighted sequencing, and (2) the region with the largest intracerebral cluster of HH-connectivity by SL, selecting the highest intra-cluster connectivity by SL (patient example in **Figure 1**, all patients in **Supplementary Table 1**). The choice and size of the SL-guided SOZs were determined by the constraints of (1) the HH size, (2) the region within the ablated portion of the HH that preoperatively had the largest and highest activation, and (3) highest correlation to the regions outside the HH in pZ pattern demonstrated by SL. The size of the pZ was primarily selected for regional grey matter thickness within this peak-appearing connectivity region, or otherwise, region of cohesive and high connectivity determined by expert review and known to be a frequent region of propagation by prior studies, and primarily the individual’s SL results (Boerwinkle et al., 2018a; Boerwinkle et al., 2016; Murta et al., 2012; Usami et al., 2016). The pZ was either drawn manually or an AAL3 hippocampus mask. Time series extraction from the SOZ and pZ were performed through a general linear model (GLM) of the rs-fMRI data. Given the prior noise correction, no additional noise correction regressors were used. Both rs-fMRI runs were entered as the same session. SPM concatenate (Casanova et al., 2007) was used to specify the boundary between the runs to correct for intensity differences between sessions. From the resulting SPM, a time series for each SOZ and pZ were extracted using the first eigenvector, as prior (Sussman et al., 2021).

**Figure 1.**
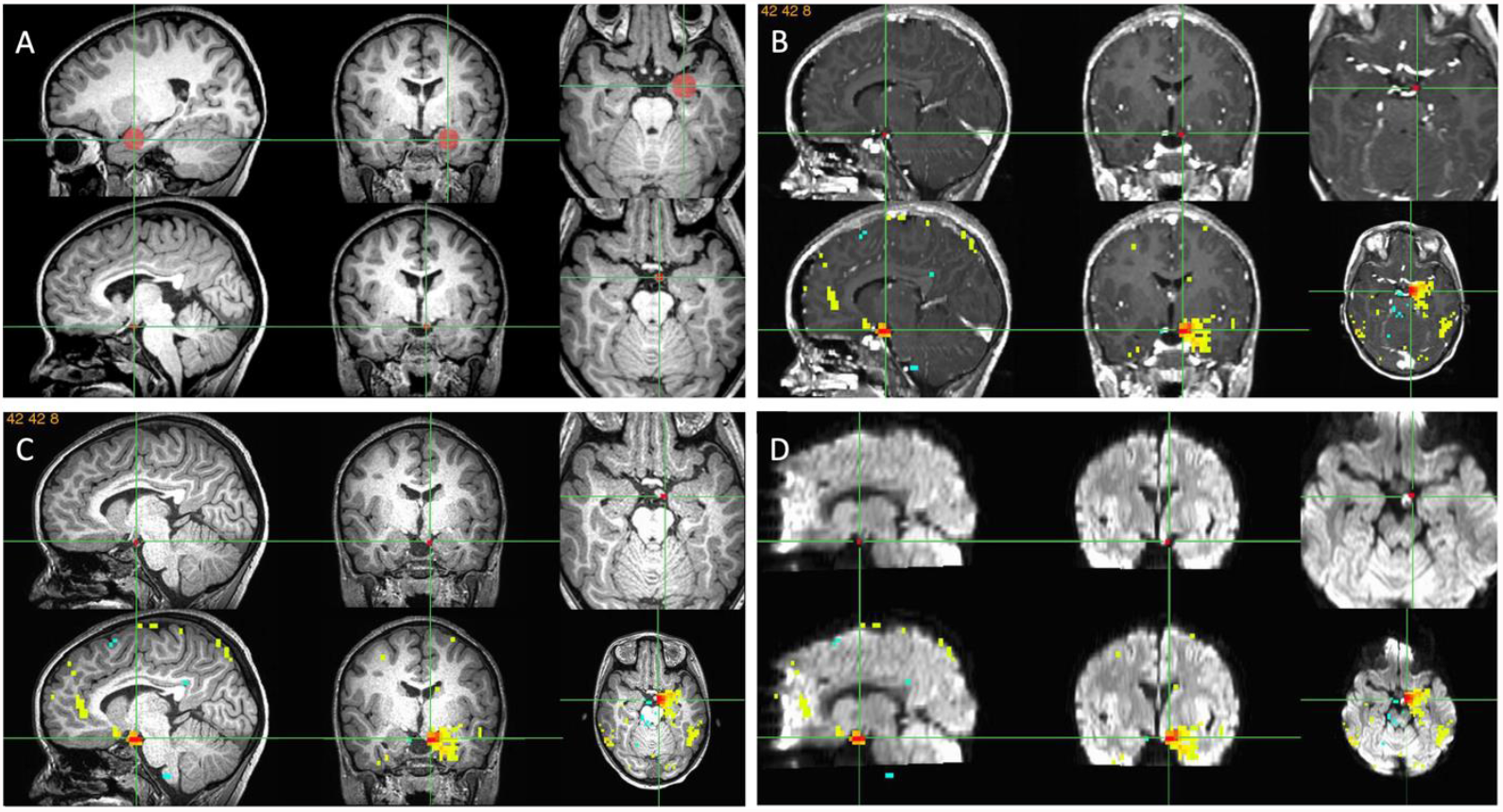
Example of ROI of the SOZ and pZ selection. Each set of six images is divided into two rows of sagittal, coronal, and axial images. **A**. Pre-operative T1W wherein row 1 is a left hippocampal pZ (red circle), and in row 2 is the SOZ within the HH, which is one voxel large. **B-D**. Post-operative post-contrast T1W, pre-operative T1W, and post-operative diffusion weighed images, respectively showing in row 1 same SOZ as in A, and row 2 includes the SL connectivity to the SOZ. These views allow visualization of the regions of the SOZ destroyed as either the post-contrast or the diffusion were positive in SOZ location and with connectivity to the pZ ROI selected. Abbreviations: ROI: region of interest; pZ: propagation zone; SOZ: seizure onset zone; SL: SearchLight.

#### 2.2.5 DCM

##### 2.2.5.1 Model Estimation

In overview, cross-spectral DCM (Friston et al., 2014a) was used to model the regional time course signals, in a fully-connected model, similar to previously described (Sussman et al., 2021). Cross-spectral DCM estimates parameters of auto- and cross-spectrum through multivariate auto-regression models of BOLD data generating estimated spectrums. As such, a two-node model was specified per case and steady-state spectral amplitude and phase representations of each node’s (ROI) first principle component activity were obtained through Fourier transform. Variational Bayesian inversion was used to fit the differential equation connectivity model (Friston et al., 2003; Friston et al., 2014b). Inversion involves fitting the DCM to maximize the likelihood of the model under prior parameter specifications. Model-fitting estimates parameters that describe the amplitude by frequency-spectral representation for each region. Each region’s local spectrum was modeled as a power law distribution with an amplitude and scale, the latter indicating the frequency by amplitude slope. Effective connectivity was derived through estimating the same parameters through frequency cross-spectrum between regions.

##### 2.2.5.2 Single Subject Model Comparison

Our aim of identifying connections in this HH network was achieved by a Bayesian model reduction (BMR) and averaging approach (BMA). Specifically, we estimated the probability that every single model generated the data through its Bayesian furnished model evidence of the fully-connected model (BMR). The model evidence was then used to weight the probability of each connectivity parameter in each fit model. These parameters were then averaged across all models to determine the statistically optimal estimate of local neural and regional connectivity effects (BMA). This approach allowed us to furnish a statistical reliability measure of estimated connectivity parameters for each tested model within a single subject, circumventing the need to compare models across subjects. A threshold of 0.9 was used for the posterior probability of each parameter for further analysis.

##### 2.2.5.3 Group Model Estimation and Comparison

In addition to single-subject model comparison, we planned a group Parametric Empirical Bayes (PEB) analysis to perform group-based model reduction and comparison to identify mean group effects as well as investigate the group effects of Engel class outcome and age on parameter estimate size. For this, the fully-connected individual DCMs were estimated, but then exhaustive Bayesian model reduction, comparison, and averaging were performed through a PEB paradigm (Friston et al., 2016; Friston et al., 2015; Zeidman et al., 2019). Exhaustive BMR was used because it is a data-driven strategy in an otherwise hypothesis driven technique. In the model, Engel class 1a outcome were coded as 1 and Engel class 1b outcome were coded as -1. The covariates were mean-centered and entered in the following order: Engel class outcome, age. Covariate effects of interest (posterior probability > 0.95) were entered into a leave-one-out (LOO) cross-validation analysis to examine their predictive ability. For discussion purposes, we focused on parameters with a posterior probability greater than 0.95 (free energy).

### 2.3 Statistical Analyses

#### 2.3.1 Demographics and comparison of single subject model results

Baseline demographics and clinical factors were summarized using count and percent for categorical variables and the mean and standard deviation for quantitative measures. Descriptive SOZ and pZ locations were quantified.

Sensitivity, accuracy, positive predictive value were determined of Directionality’s identification of the SOZ and propagation zones in individuals’ models from the single subject model estimation and comparison. Due to study design, there were no true negatives, thus specificity and negative predictive value are not determined. The proportion of agreement between Directionality and surgical outcome with 95% binomial exact confidence interval (CI) was calculated by node-to-node connection. The agreement of Directionality and surgical outcome was assessed using the prevalence-adjusted bias-adjusted kappa (PABAK)(Byrt et al., 1993) instead of the traditional kappa since prevalence was expected as study design did not allow for true negatives. A groupwise effective connectivity PEB analysis was performed as described above.

#### 2.3.2 Bias

To avoid bias we also explored whether PEB results were likely related to additional clinical, demographic, or analysis-related (e.g. ROI size) variables, we also planned a series of tests as follows: To further explore if PEB results are were explainable by clinical and demographic characteristics, we correlated age with SOZ size, pZ size, pre-operative seizure frequency, and age. Also, to ensure that ROI size was not associated with DCM outcomes, we correlated the ROI sizes of the SOZ and pZ with baseline (A matrix) connectivity parameters from the PEB analysis. Finally, although it was not included in the PEB model, we examined whether sex was associated with baseline parameter estimates from the PEB, as well as pre-to-post operative seizure percent improvement. Independent sample comparisons of continuous variables were tested using two-sided t-tests or Mann-Whitney tests in cases of violations of assumptions of normal data. Correlations were tested using two-sided Pearson’s r tests. Bonferroni corrections were used. All confounder analysis tests were performed with JASP Statistics v0.16. (JASP Team, 2021)

## 3. Results

### 3.1 Descriptive results

Individual patient clinical characteristics are summarized in **Table 1** and expanded on in **Supplementary Table 2**. Of the 28 patients, the mean age was 7.5 years (standard deviation (SD) of 4.8 years, range of 2.1-18.2 years), with a male to female ratio of 18:10. The pZs were both cortical and subcortical (28 cortical:4 subcortical), and in greater proportion in the frontal and temporal lobes (**Table 1**). The median size of the HH mask was one 3×3×3.4mm voxel [interquartile range (IQR): 1-1; range: 1-11] and the median size of the pZ mask was sixty-five 3×3×3.4mm voxels [IQR: 8-181.3; range: 1-227].

**Table 1.** Demographics and pZ Location

| N=28 |  |  |
| --- | --- | --- |
| Sex (M:F) | 18M : 10F |  |
| Handedness | 14R : 7L : 7ND |  |
| Age (Mean, SD) | (7.49, 4.82) |  |
| pZ | Left/Right/Bilateral | Total |
| Hippocampus | 3/4/0 | 7 |
| Frontal | 3/4/0 | 7 |
| Temporal | 3/3/0 | 6 |
| Parietal | 1/0/0 | 1 |
| Occipital | 0/1/0 | 1 |
| Cingulate | 0/0/1 | 1 |
| Basal ganglia | 3/0/0 | 3 |
| Thalamus | 1/0/0 | 1 |
| Brainstem | 1/0/0 | 1 |
| **TOTAL** | **15/12/1** | **28** |

Sex: M – male, F – female; Handedness: R – right, L – left, ND – non-dominant; SD – standard deviation; pZ – propagation zone. Handedness was determined by epileptologists’ documentation in patient’s medical record.

### 3.2 Comparisons with individually estimated and reduced models

Results from the individual DCM estimation, model reduction, and Bayesian model averaging for an example subject are shown in **Figure 2**. Individual model results for all subjects can be found in **Supplementary Table 1**. When considering the SOZ effective connectivity, 3.6% had no significant connection (**Table 2**). Of the remainder with significant connections, 96% and 3.7% were in the expected and reversed directions, respectively (**Table 2**). Positive and negative polarity signals were equivalent as expected between the SOZ and pZ (**Table 2**). By connection, considering both positive and negative, Directionality had sensitivity of 96%, accuracy of 93%, and positive predictive value of 96%. The observed agreement between Directionality and surgical outcome was 92.9% (N=52/56; 95% CI: 0.83-0.98). The PABAK estimate was 0.86 (95% CI: 0.65-0.96), indicating substantial agreement between Directionality and surgical outcome.

**Figure 2.**
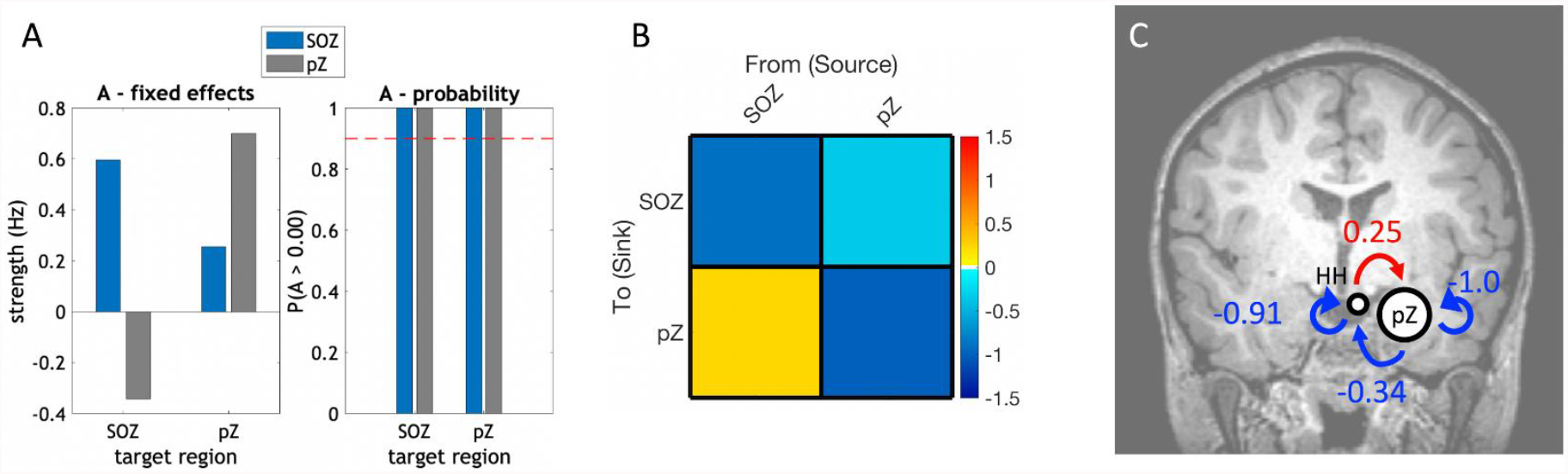
Individual DCM outputs. These outputs were calculated for each patient. **A**. Parameter estimates and their posterior probabilities of the fully specified DCM A matrix (intrinsic connectivity) after inversion using cross-spectral DCM. Target region is listed on the x-axis; connections from the SOZ (HH) are in blue and connections from the pZ are in grey. The left graph is the effect sizes, self-connections are shown in the log scaling value used to stabilize the model (conversion to Hz is -0.5 * exp(A)). The right bar chart shows the posterior probabilities of each estimated parameter. **B**. Adjacency matrix of the A matrix parameter estimates after optimization (exhaustive BMR followed by BMA). This matrix is thresholded to only include connections with a posterior probability greater than 0.9. Self-connections have been converted from the log-scaling value to Hz. **C**. The values from panel C shown as a diagram superimposed on the patient’s coronal T1W image. Abbreviations: DCM: dynamic causal modeling; ROI: region of interest; pZ: propagation zone; SOZ: seizure onset zone; SL: SearchLight; HH: hypothalamic hamartoma.

**Table 2.** Directionality Results and Signal Characterization

|  | Positive/<br>Excitatory | Negative/<br>Inhibitory | No<br>Significant<br>Connection | % |
| --- | --- | --- | --- | --- |
| SOZ→pZ (n) | 26 | 1 | 1 |  |
| pZ→SOZ (n) | 1 | 26 | 1 |  |
| % Expected of those<br>with detected signal |  |  |  | 96 |
| Signal reversal (%) of<br>those with signal |  |  |  | 3.7 |
| Lack of signal (%) |  |  |  | 3.6 |

| Predicted<br>Condition | True Condition |  |
| --- | --- | --- |
|  | Concordant Signal<br>Direction | Discordant Signal<br>Direction |
| Nodal<br>relationship<br>detected | SOZ→pZ pos (26) | SOZ→pZ neg (1) |
|  | pZ→SOZ neg (26)<br>52 TP | pZ→SOZ pos (1)<br>2 FP |
| Nodal<br>relationship not<br>detected | SOZ→pZ no sig (1) |  |
|  | pZ→SOZ no sig (1)<br>2 FN | 0 TN |

Abbreviations: n: number; neg: negative; pos: positive; pZ: propagation zone; SOZ: seizure onset zone; TP: true positive; FP: false positive; FN: false negative; TN: true negative; sig: significant signal

### 3.3 Parametric Empirical Bayes Group Analysis Results

The BMA group model is shown in **Figure 3A**. The connection from the SOZ to the pZ was excitatory and the connection from the pZ→SOZ was inhibitory. The SOZ self-connection was also present (inhibitory). The pZ self-connection was pruned during BMR.

**Figure 3.**
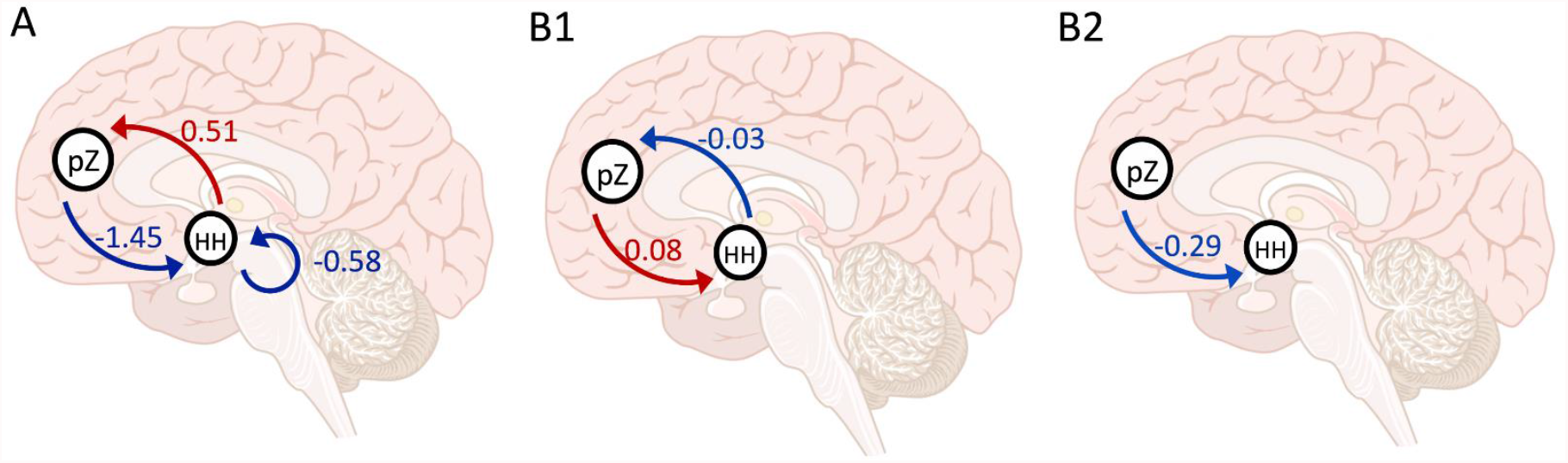
**A**. Group baseline connections between the SOZ and pZ thresholded at 0.95 posterior probability after exhaustive Bayesian Model Reduction (BMR) and Bayesian Model Averaging (BMA). The location of the pZ varied by subject. Red arrows and values are excitatory modulation, blue arrows and values are inhibitory modulation. **B**. Group connections associated with Engel outcome and age thresholded at 0.95 posterior probability. Blue arrow/value indicates a negative correlation and a red arrow/value indicates a positive correlation. **B1**. Engel class Ia outcomes were coded with a higher value than Engel class Ib outcomes, thus, the blue arrow in Panel A indicates that patients with better one year outcomes (Engel class Ia) showed more inhibitory modulation from the pZ to SOZ. **B2**. Older patients show less inhibition from the pZ to SOZ and less excitation from the SOZ to pZ, thus, overall, absolute strength of these parameter effect sizes decreased with age

Regarding behavior correlations, Engel class outcome was associated with signal strength ***<u>from the pZ to the SOZ</u>*** (**Figure 3B**). Patients with a better Engel class outcome (1a) showed greater inhibitory signal from the ROI to the SOZ **(Figure 3B1)**. However, this difference, however, was not significant with LOO cross-validation (r(26)=-0.07, *P*=0.65; **Supplementary Figure 1A**). Age was also associated with between-node connections; age was positively correlated with signal from the pZ→SOZ and negatively correlated with signal from the SOZ→pZ (Figure 3B2). Neither of these correlations were significant with LOO cross-validation r(26)=0.08, *P*=0.33 and r(26)=0.21, *P*=0.14, respectively (**Supplementary Figure 1B-C**). Self-connections were not related to Engel outcome or age.

### 3.4 Bias comparisons results

Mann-Whitney test results were not significant for differences between Engel outcomes and patient variable of pre-operative seizure rate, SOZ, and pZ size (**Supplementary Table 3**). Further, age at scan was not correlated with SOZ size, pZ size, seizure frequency, nor seizure frequency improvement (%), nor were SOZ or pZ size correlated with A matrix parameter estimates from the PEB analysis (**Supplementary Table 4**). Student t-tests were not significant for sex differences for A matrix parameter estimates from the PEB analysis and a Mann-Whitney test was not significant for sex differences in seizure frequency improvement (%) **(Supplementary Table 5)**.

## 4. Discussion

This is the first reported DCM analysis of rs-fMRI, performed independent of electrophysiological data, to demonstrate the capacity of RSEC alone to differentiate SOZ from the region of propagation with high accuracy, sensitivity, and positive predictive value. This is also the first study to demonstrate that cross-spectral DCM of rs-fMRI yields the excitatory signal direction from SOZ to a region of propagation and inhibitory signal from the region of propagation back to the SOZ, on both in the individual and group level. Before application to the more heterogeneously localized SOZ of the general DRE population, it was necessary to perform this analysis in such a homogenous and well established SOZ location of the HH to understand the potential of this tool for narrowing ROI for SOZ location determination. Importantly, polarity of SOZ and pZ connections were *<u>consistent in both individual and group analyses</u>*, increasing confidence in individual application, similar to prior RSFC (Boerwinkle et al., 2019a; Boerwinkle et al., 2018a; Boerwinkle et al., 2017; Boerwinkle et al., 2016) and RSEC work (Vaudano et al., 2021).

Our hypothesis that, from rs-fMRI alone, intrinsic modulation from the SOZ is excitatory and modulation from the pZ is inhibitory is supported by our results. Unlike previous studies, we did not time lock or model around ictal onset or known periods of interictal discharge (Daunizeau et al., 2012; Hamandi et al., 2008; Klamer et al., 2018; Klamer et al., 2015; Murta et al., 2012; Vaudano et al., 2013; Vaudano et al., 2009; Vaudano et al., 2021; Warren et al., 2019). Instead, we assumed, that in DRE, markers of epileptic signal are present in resting-state networks regardless of known or unknown ictal and interictal activity at time of data capture and without time-locking to ictal/interictal activity. This has been supported in previous studies using RSFC to reliably identify the (static) spatial locations of epileptogenic networks (but not the direction of signal propagation) between regions in these networks without time-locking to epileptogenic activity (Boerwinkle et al., 2018a; Boerwinkle et al., 2017; Boerwinkle et al., 2018b; Boerwinkle et al., 2016). We expanded on previous findings to show that information about direction of epileptogenic signal is also present in event-agnostic rs-fMRI signal.

While it is possible to use methodologies to know whether patients were seizing in the scanner or would have shown concurrent epileptiform activity, the consistency of the results herein is a strength that implies that, in this population, such knowledge may not be necessary in order to model seizure propagation direction but requires further study in a larger population of these patients who had poor outcomes and see if the model remains consistent. In fact, necessitating EEG-fMRI and capture of epileptiform event in scanner can reduce the overall clinical yield of fMRI-based EC techniques in epilepsy network mapping. Using EEG-fMRI to guide DCM, Vaudano et al. (2021) found that all 4 patients whose DCM indicated SOZ was concordant with the clinical SOZ and surgically-targeted had good clinical outcomes. However, this subsample represented only a 14.3% (4/28) surgically-validated clinical yield of all patients who underwent EEG-fMRI and would likely benefit from an EC analysis, compared to a surgically-validated clinical yield of 81.3% (26/32) in the current study (see **Supplementary Table 6**), a Fischer exact test found this difference statistically significant (*P*<0.001). A broad summary of methodological differences between the current study and prior studies is detailed in **Table 3**.

**Table 3.** Comparison of Methods in Prior and Current Work

| Method Differences | Prior Work (EEG-fMRI) | Current Study (rs-fMRI) | Comment |
| --- | --- | --- | --- |
| ROI selection | - Requires simultaneous EEG-fMRI, clinically cumbersome<br>- Requires epileptic discharge (interictal/ictal epileptic activity)<br>- ROI specification guided by identifying BOLD response associated with epileptogenic activity<br>- ROI specification has also incorporated information from MEG (Klamer et al., 2015) and tractography (Hamandi et al., 2008) | - Unimodal rs-fMRI, EEG not needed<br>- Static RSFC (ICA, SL), guides ROI specification as cross-spectral DCM relies on generalized measures of RSFC (Friston et al., 2014a) making RSFC an appropriate basis for ROI selection<br>- No requirement for capture of epileptiform activity | - Advancement *is independence from capturing epileptiform events* and cumbersome equipment, which increases yield of study from 14.3% to 81.6% (**Supplementary Table 6**) with **sensitivity of current study 96%**<br>- Both require ROI selection method |
| Assumptions about epileptogenic drivers from subtype of timed events | - GPFA (Warren et al., 2019)<br>- IED (Hamandi et al., 2008; Klamer et al., 2018; Vaudano et al., 2013; Vaudano et al., 2009; Vaudano et al., 2021)<br>- ictal (David et al., 2008; Klamer et al., 2015; Murta et al., 2012) | - None | - EEG-fMRI assumes captured events indicate accurate direction of signal to identify SOZ, whereas rs-fMRI alone assumes excitation from SOZ and inhibition from pZ will be consistent and accurate to determine SOZ |
| DCM method subtype | - Deterministic timeseries (David et al., 2008; Hamandi et al., 2008; Klamer et al., 2018; Klamer et al., 2015; Murta et al., 2012; Vaudano et al., 2013; Vaudano et al., 2009; Vaudano et al., 2021; Warren et al., 2019)<br>- Stochastic, with event specification (Daunizeau et al., 2012) | - Cross-spectral | - Advancement is speed of analysis (model estimation) |
| DCM matrices specified | - A matrix (baseline (intrinsic) connectivity)<br>- B matrix (modulatory (context/edge) inputs)<br>- C matrix (driving (direct/node) inputs)<br>- D matrix (intrinsic gating) | - A matrix (baseline (intrinsic) connectivity) | - Matrices are dependent on inputs available, B-D matrices are related to extrinsic timed events, whereas the A matrix is from intrinsic (nonevent related) brain activity |
| Information that can be gleaned from each DCM method subtype | - Able to explicitly model SOZ by specifying direct/driving input to nodes and compare likelihood of driving input location (e.g., whether driving input during ictal/interictal event is to node A or B) and explicitly model modulation of connection during ictal/interictal event (e.g. whether modulation is to the A→B connection or the B→A connection)<br>- EEG - BOLD events can be compared | - Can only use A matrix (intrinsic connections), does not test hypotheses about specific timed modulation of connections or direct input to nodes | - EEG-fMRI allows observation of timed signal transmission to determine node order, however *rs-fMRI alone had 93% accuracy* |
| Model selection | - Generally, hypothesis driven model specification with FFX comparison<br>- BPA | - Exhaustive BMR with BMA | - Advancement is data-driven model, wherein no assumption of relationship between nodes is made. BMR is also less a less computationally intensive nested model estimation and comparison scheme than separately estimating each model |
| Evaluation of excitation in SOZ identification | - Yes, reported single patient validated with iEEG (Vaudano et al., 2013), Single patient with EEG-fMRI (Hamandi et al., 2008; Murta et al., 2012) | - Yes, 96% of HH SOZ with excitation to pZ | - Corroboratory, but prior method with reduced SOZ confirmed detection comparatively |
| Evaluation of inhibition in SOZ identification | - Reported but not interpreted as meaningful (Vaudano et al., 2013) | - Yes, identifies inhibition in A matrix as most common 'feedback' from pZ to the SOZ | - Advancement is further check on SOZ identification by differentiating inhibition as coming from the pZ as opposed to the SOZ |
| Validated by surgical outcomes | - Largest study N=10, 6 with surgical outcome (Vaudano et al., 2021) | - Current study (N=28) | - Most studies are without comparison to surgical outcomes |

Abbreviations: SL: SearchLight; ICA: independent components analysis; IED: interictal epileptic discharge; EEG: electroencephalography; FFX: fixed effects; GPFA: generalized paroxysmal fast activity; BPA: Bayesian Parameter Averaging; BMA: Bayesian Model Averaging; RSFC: resting state functional connectivity

Our current results show some differences in terms of polarity from prior studies. For example, we show that the connection from pZ→SOZ is overwhelmingly negative; however, pZ→SOZ connections in prior event-based DCM studies in have been mostly reported positive (Hamandi et al., 2008; Murta et al., 2012; Vaudano et al., 2013) with one study showing a mix (Vaudano et al., 2013). Thus, our results show greater consistency of A matrix connection polarities. This difference may be due to the number of nodes modeled, differences in ROI specification, patient effects (all of our patients had Engel class I outcomes), or potentially more likely – our model did not include direct inputs or modulatory influences during captured explicit epileptiform activity and used cross-spectra (second-order statistic) instead of time-varying fluctuations (time series). As such, we also assume that the state of the relationship between nodes in the current study is modelling the basal state of the relationship between SOZ and pZ without placing temporal emphasis on ictal or inter-ictal events. This may be different from their relationship during these events, thus contributing to differences seen in our analysis. However, the differences could also be related to volatility of nodal relationship during epileptiform activity, as nodal strength and polarity inconsistencies in prior EEG-fMRI are prevalent (David et al., 2008; Murta et al., 2012; Vaudano et al., 2013), whereas Directionality was highly consistent.

Also consistent for Directionality was network architecture. Bi-directional connections between SOZ and pZ had overwhelmingly high probability, which is not dissimilar previous time-locked DCM studies that compared models with and without bidirectional connections along propagation pathways (Murta et al., 2012; Warren et al., 2019). Thus, our results imply that both SOZ→pZ and pZ→SOZ connections in baseline epileptogenic networks are relevant and, in a two-node model, can be identified with exhaustive BMR rather than specifying models for BMR.

Both of the groupwise covariates evaluated were revealing. Engel class Ia versus Ib outcomes showed greater inhibitory signal from pZ→SOZ. Assuming that inhibitory pZ→SOZ signal is suppressive, then this effect may indicate that not only is SOZ ablation needed for good outcome, but also a stronger pre-operative pZ inhibition encourages possible residual SOZ activity to remain below the seizure threshold. ***Notably, the potential of measuring a precision network-dependent factor of pZ inhibition to predict seizure outcome from rs-fMRI alone is novel***. Further work is indicated, as despite very strong posterior evidence, LOO cross-validation results were not significant. Likely, broader Engel class I-IV outcomes would clarify if the pZ inhibition strength is associated with surgical outcomes.

Absolute signal strength emitted from SOZ and pZ was smaller as age increased. However, these associations did not survive LOO cross-validation, limiting current predicative power of directional signal strength. Decreasing excitation from SOZ→pZ with age is novel, though counterintuitive, given that strength of aberrant signal is expected to increase with disease length. However, recent DCM of epileptiform propagation pathways of Lennox-Gastaut Syndrome also showed reduction with age (Warren et al., 2019). This being noted, the strength difference did not have an effect on SOZ detection yield in this group. Further, the strength difference over age is relatively small, thus not expected to have an effect on SOZ detection in other DRE populations.

Engel outcomes and pre-operative seizure frequency and ROI size were unrelated. Further, age, ROI size, and pre-operative seizure frequency were unrelated to each other. The lack of relationship between surgical outcome and pre-operative seizure frequency and ROI, as well as between variables, make it unlikely that these factors biased Directionality. RSEC strength correlated with Engel outcome, but not seizure frequency nor reduction. Thus, severity of epilepsy also does not appear to influence the applicability of Directionality.

### 4.1 Limitations

Because all patients included by study design had good outcomes, it is not known how Directionality will perform in the broader outcome nor heterogenous DRE cause population. Although this study used a homogenous group and only two nodes, it provides a foundational basis for expansion to more complex patients and models. Our method of ROI selection does not preclude the possibility of additional pZs, that any given pZ is the ‘first’ pZ from the HH-SOZ, nor a ‘longer’ propagation pathway. Since patients with HH generally do not receive iEEG for surgical planning, it is not possible to determine the absolute truth of the pZ. However, since all HH-SOZs were selected to be within both the area of surgical destruction that lead to increased or total seizure freedom and within a location determined by SL to have high RSFC outside of the HH we are confident that SOZ ROI selection is accurate.

### 4.2 Future Directions

Noninvasive characterization of regional excitation/inhibition relationships may prove clinically impactful in other neurological disorders amenable to precision-based network-targeted interventions such as in the broader DRE population, and possibly movement (Sussman et al., 2021), neurodegenerative, and neuro-psychiatric disorders. The current models depict seizure propagation pathways in two node models where the SOZ is known. Future directions should also investigate patients with suboptimal surgical outcomes as well as other epilepsies with other etiologies and models with more nodes.

## 5. Conclusions

This study demonstrates the high accuracy (93%) and sensitivity (96%) of Directionality to identify the origin of excitatory and inhibitory signal between SOZ and the hypothesized pZ in children with HH-DRE. Directionality can be performed from 20 minutes of rs-fMRI data and does not require the additional cumbersome or expensive procedures, personnel, or equipment needed for EEG-fMRI. It also benefits from a lack of need to capture epileptiform activity and ultimately leads to faster results due to greater simplicity of solution – which may also lead to a higher clinical yield. This method validation study in a homogenous population with known SOZ location and surgical outcomes may be helpful in narrowing the SOZ in regions of suspicion for epileptogenicity and may ultimately decrease dependency on iEEG in other DRE populations and other neurological disorders.

## Data Availability

The data made available under institutional IRB approval that supports the findings of this study are available in the supplementary material of this article.

## CRediT Author Statement

**Varina Boerwinkle:** Conceptualization, Methodology, Validation, Formal Analysis, Investigation, Resources, Writing – Original Draft, Writing – Review & Editing, Visualization, Supervision, Project Administration; **Bethany Sussman:** Conceptualization, Methodology, Software, Validation, Formal Analysis, Investigation, Writing – Original Draft, Writing – Review & Editing, Visualization; **Sarah Wyckoff:** Conceptualization, Investigation, Formal Analysis, Data Curation, Writing – Review & Editing; **Iliana Manjón:** Visualization, Writing – Review & Editing; **Justin Fine:** Software, Writing – Review & Editing; **P. David Adelson:** Writing – Review & Editing

## ^1^Abbreviations

BMA: Bayesian Model Averaging
BMR: Bayesian Model Reduction
BOLD: blood oxygen level dependent
CAT12: Computational Anatomy Toolbox version 12
DCM: dynamic causal modeling
DRE: drug resistant epilepsy
EEG: electroencephalography
EEG-fMRI: simultaneous EEG and functional MRI
GLM: general linear model
HH: hypothalamic hamartoma
ICA: independent component analysis
iEEG: intracranial EEG
IED: interictal epileptogenic discharges
IED-BOLD: simultaneous EEG-fMRI detected interictal epileptogenic discharges corresponding to blood oxygen level dependent rs-fMRI signal changes
LITT: laser interstitial thermal ablation therapy
MEG: magnetoencephalography
PCA: principal components analysis
PEB: Parametric Empirical Bayes
pZ: propagation zone
ROI: region of interest
rs-fMRI: resting state functional MRI
RSFC: (static) resting state functional connectivity
RSEC: resting state effective connectivity
SL: SearchLight
SOZ: seizure onset zone

## Acknowledgements

The authors would like to acknowledge the nonprofit organization Hope for Hypothalamic Hamartomas for the dedicated advocacy and research funding needed to forward progress for rare epilepsies, and for supporting the work that has borne out to be now a well-established human epilepsy network, allowing for a homogenous DRE to pave the way for discoveries for the entire epilepsy population. We would also like the acknowledge the neurosurgeons and epileptologists who supported research in this area, Daniel J. Curry, MD, P., John F. Kerrigan, MD, and Angus A. Wilfong, MD. This work stands on your shoulders. Lastly, but most importantly, to the patients with hypothalamic hamartoma and your families, who inspire us to be better. We humbly dedicate this work to the memory of Grace Katherine Webster, our constant reminder that SUDEP is a devastating puzzle that must be solved!

