## Supplemental Figure1 for "Noninvasive Epileptogenic Signal Direction Determination by Effective Connectivity of Resting State Functional MRI - Independent of EEG"

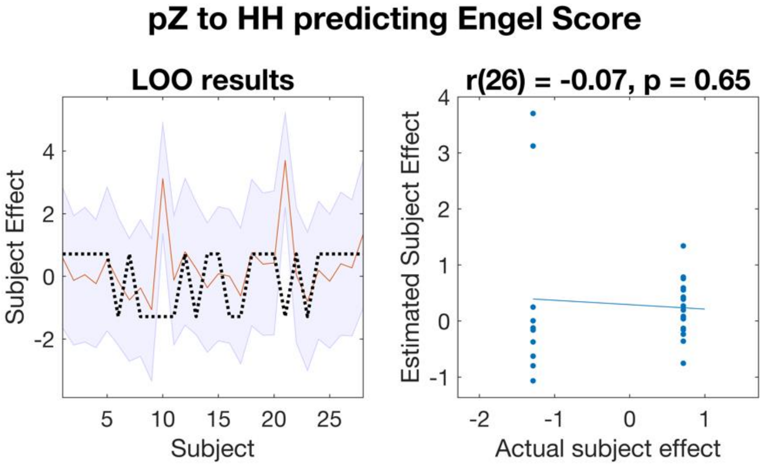


**Figure S1A.** Leave-one-out (LOO) cross-validation of pZ to HH-SOZ connection predicting Engel class outcome. LOO cross-validation is not significant. LOO cros-validation was performed by iteratively removing each subject's data and, predicting their data given the rest of the subjects' data, and then comparing their estimated value to the actual value for that subject. **Left**: Plots the out-of-samples estimate of the mean-centered subject (effect of) Engel Class outcome (black dotted line) for each subject plus variance (purple shaded area, 90% CI) and the actual group effect (red). The narrower the purple envelope, the higher the predictive degree of confidence and consistency for Engel Class outcome. The envelope is quite large, here. **Right:** Scatter plot comparing the actual subject effect to the estimated subject effect of the Engel Class outcome using an out-of-samples Pearson correlation coefficient (r). Abbreviations: pZ: propagation zone; SOZ: seizure onset zone; HH: hypothalamic hamartoma.

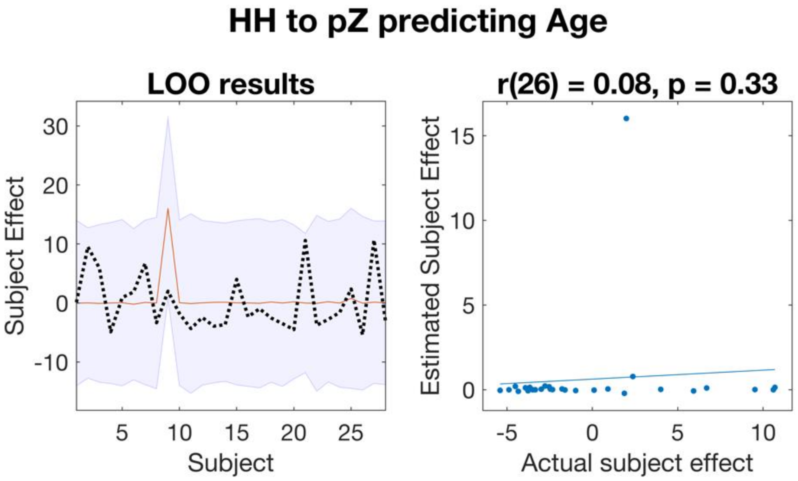


**Figure S1B.** Leave-one-out (LOO) cross-validation of HH-SOZ to pZ connection predicting age. LOO cross-validation is not significant. LOO cross-validation was performed by iteratively removing each subject's data and, predicting their data given the rest of the subjects' data, and then comparing their estimated value to the actual value for that subject. **Left**: Plots the out-of-samples estimate of the mean-centered subject (effect of) age (black dotted line) for each subject plus variance (purple shaded area, 90% CI) and the actual group effect (red). The narrower the purple envelope, the higher the predictive degree of confidence and consistency for age. The envelope is also quite large, here. **Right:** Scatter plot comparing the actual subject effect to the estimated subject effect of the Engel Class outcome using an out-of-samples Pearson correlation coefficient (r). Abbreviations: pZ: propagation zone; SOZ: seizure onset zone; HH: hypothalamic hamartoma.

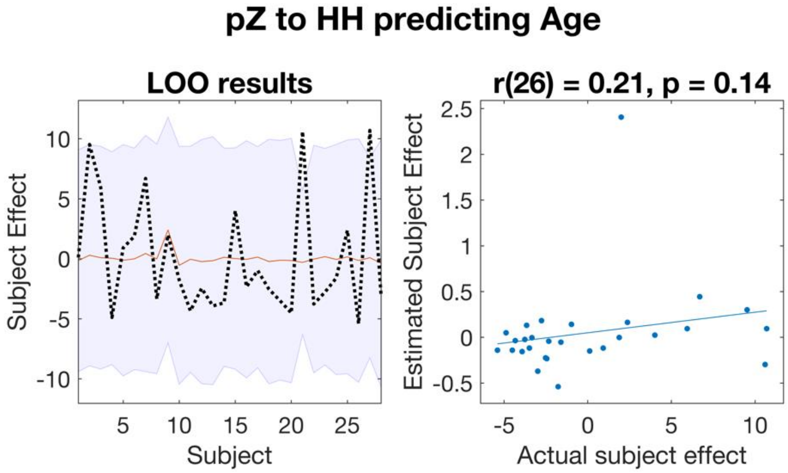


**Figure S1C.** Leave-one-out (LOO) cross-validation of pZ to HH-SOZ connection predicting age. LOO cross-validation is not significant. LOO cross-validation was performed by iteratively removing each subject's data and, predicting their data given the rest of the subjects' data, and then comparing their estimated value to the actual value for that subject. **Left**: Plots the out-of-samples estimate of the mean-centered subject (effect of) age (black dotted line) for each subject plus variance (purple shaded area, 90% CI) and the actual group effect (red). The narrower the purple envelope, the higher the predictive degree of confidence and consistency for age. The envelope is large here as well. **Right:** Scatter plot comparing the actual subject effect to the estimated subject effect of the Engel Class outcome using an out-of-samples Pearson correlation coefficient (r). Abbreviations: pZ: propagation zone; SOZ: seizure onset zone; HH: hypothalamic hamartoma.
