## Supplemental Table 1 for "Noninvasive Epileptogenic Signal Direction Determination by Effective Connectivity of Resting State Functional MRI - Independent of EEG"

**Supplementary Table 1. Patient ROIs, Pre and Post-Operative Imaging, and Directionality Result Matrices**

| # | A. Directionality HH SOZ: **sagittal, axial, and coronal T1W with HH SOZ overlayed in RED** | B. Directionality Propagation ROI: **sagittal, axial, and coronal T1W with propagation ROI overlayed in RED** | C. Pre-operative Searchlight used to select the HH SOZ and Propagation ROI for Directionality: **top row sagittal, coronal, and axial T1W with HH SOZ voxel from SL selected to be the HH SOZ for directionality based on bottom row of images with show the region of connectivity in the rest of the brain to which the voxel had greatest connectivity. Note aliased noise outside of brain occurs.** | D. Post-operative Images Used to verify Ablation of the HH SOZ for Directionality**: Diffusion weighted, and post-contrast images taken imme after ablation. Patient 5-28 with same pre-op SL results overlayed direct post-op imaging for direct verification.** | E. Directionality Result Matrix. **Note ROI location name became hard coded thus not necessarily reflecting actual ROI location description. How to read the matrix: example patient 1 – From HH to HH is white, therefore no significant connection. Clust to HH is dark blue thus strong inhibition. HH to Clust is yellow-orange thus positive. Clust to Clust is light blue thus mild inhibition.** |
|---|---|---|---|---|---|
| 1 | 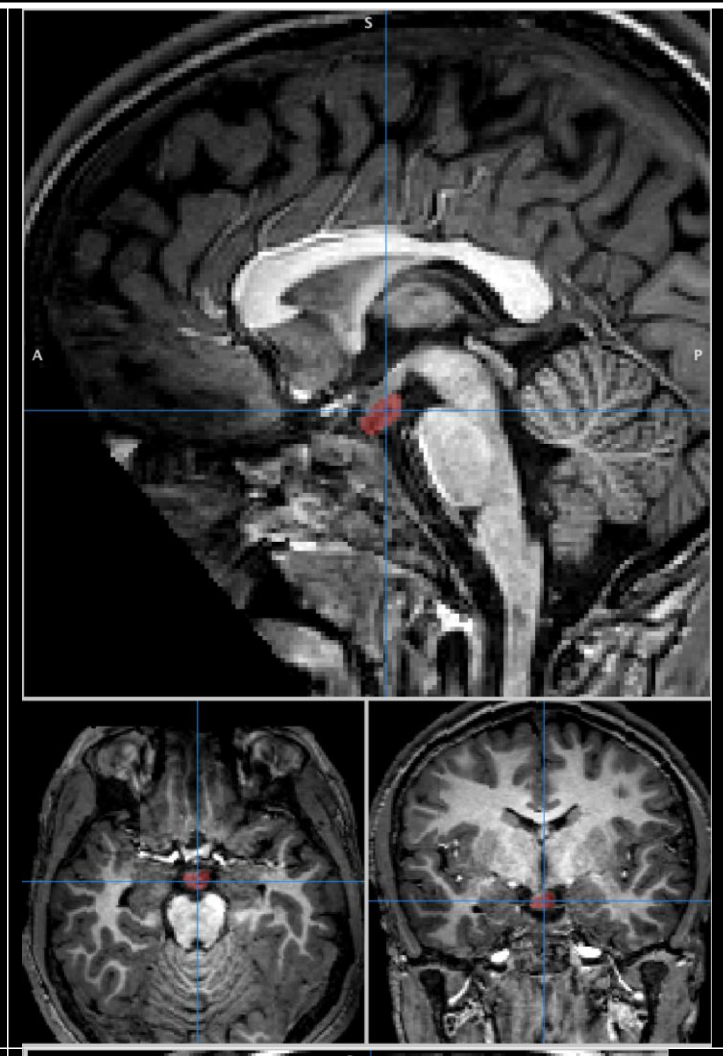 | 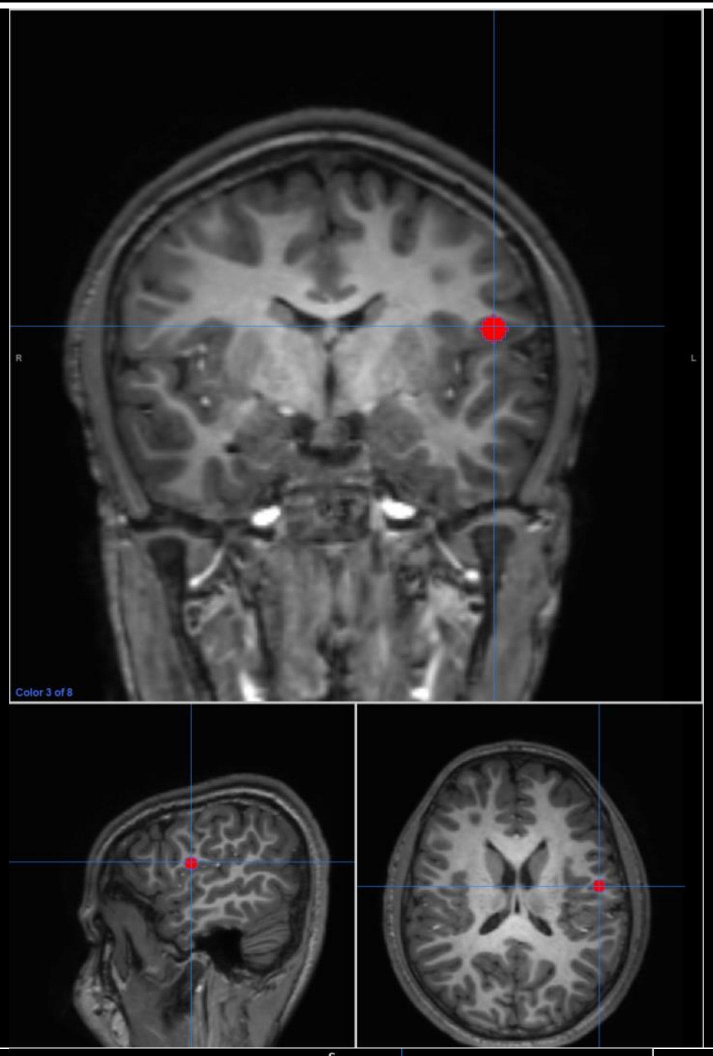 | 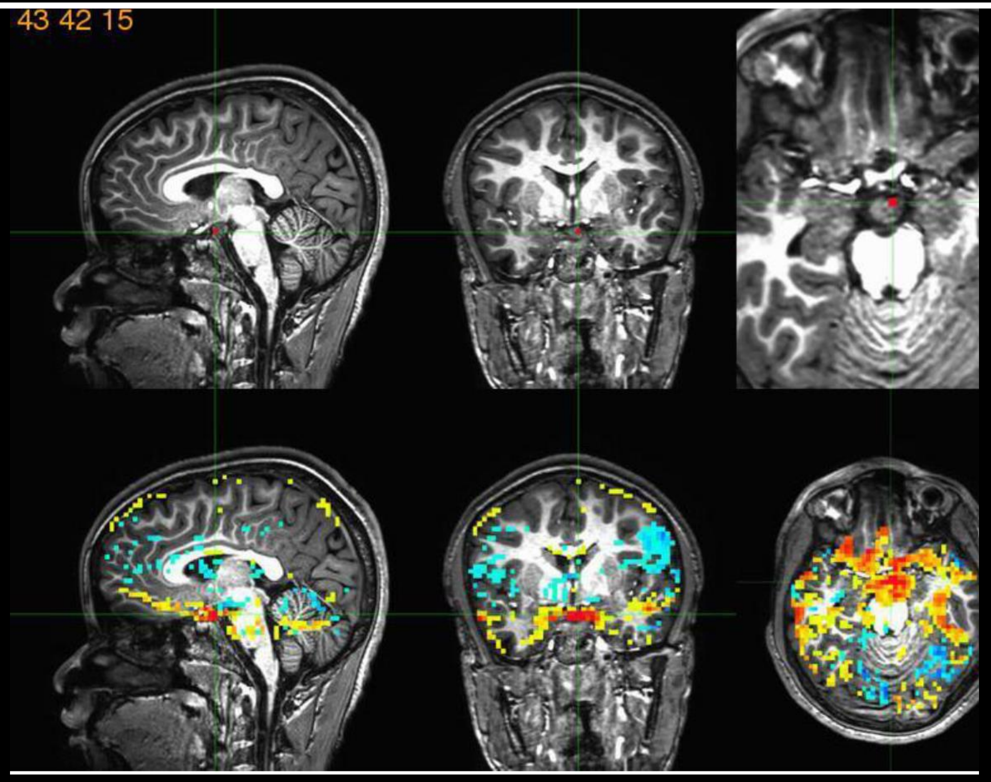  | 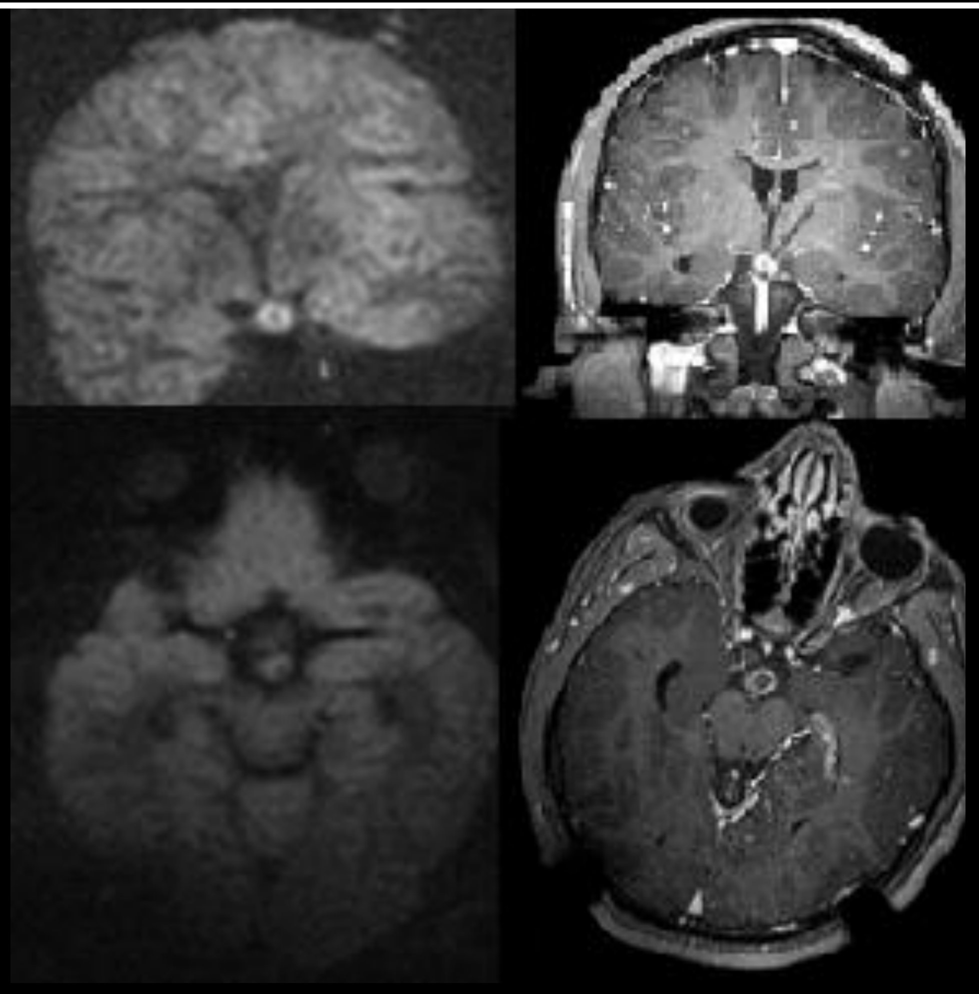 | 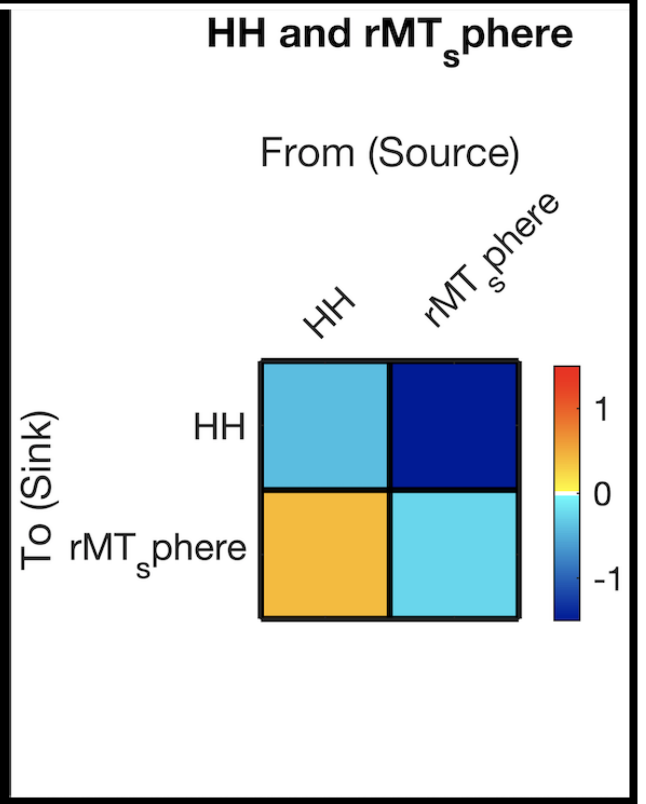  |
| 2 | 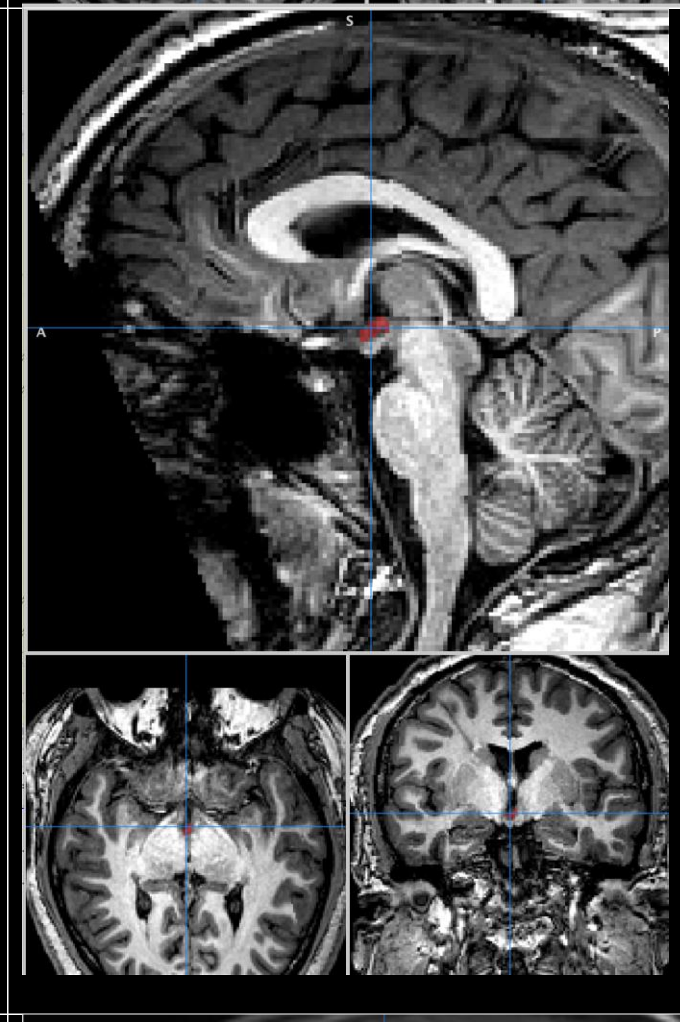 | 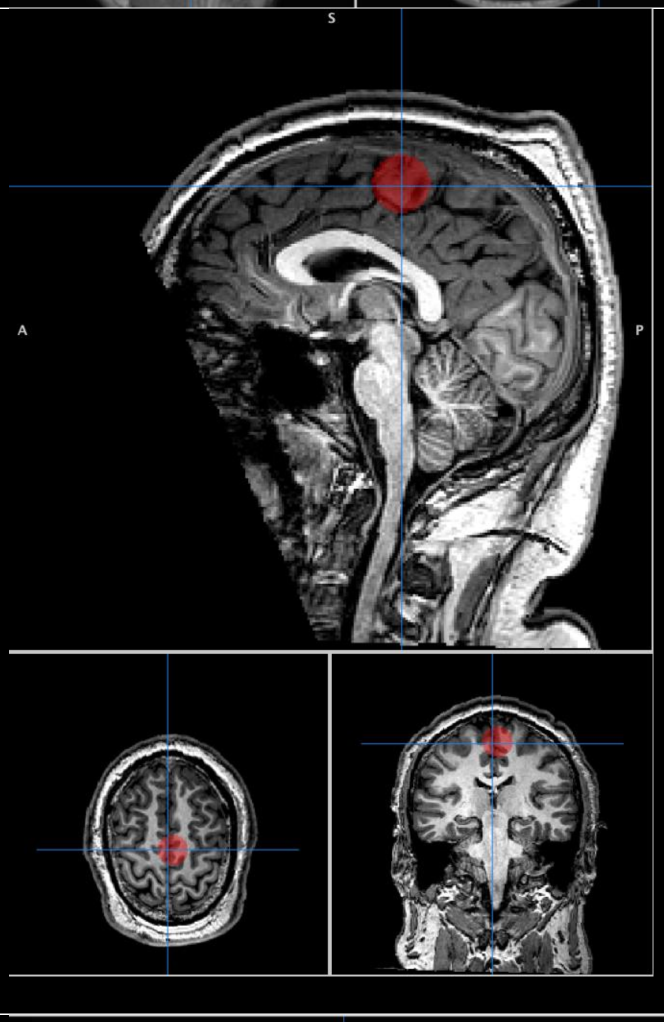 | 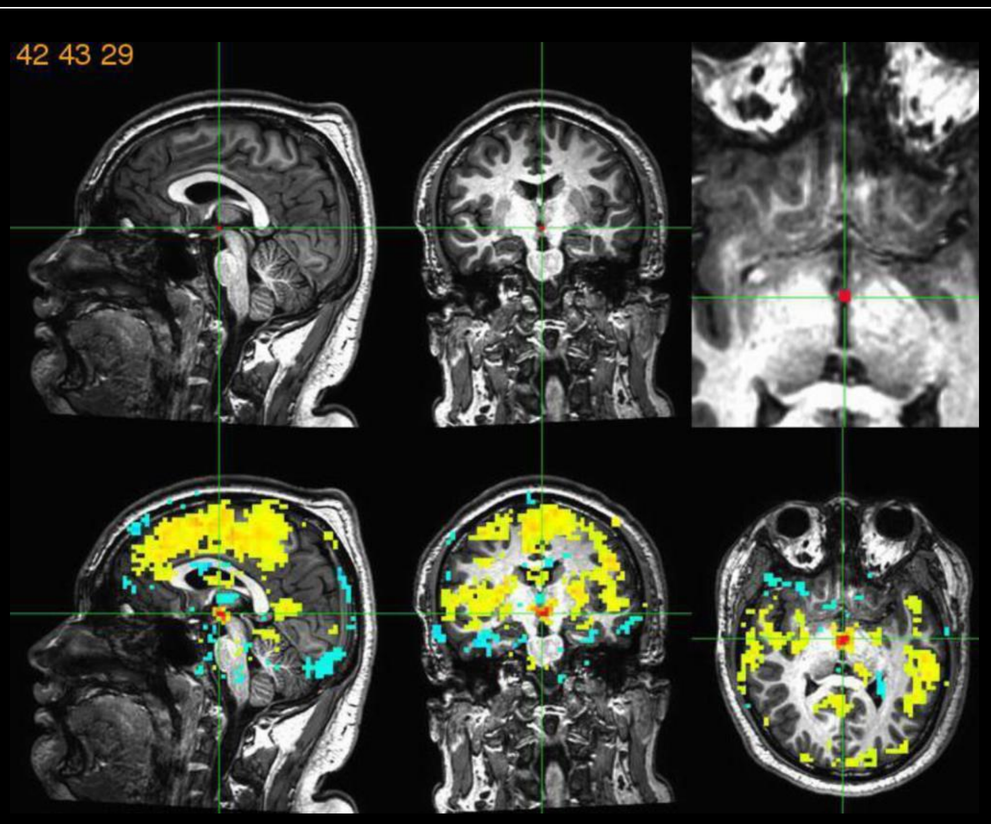  | 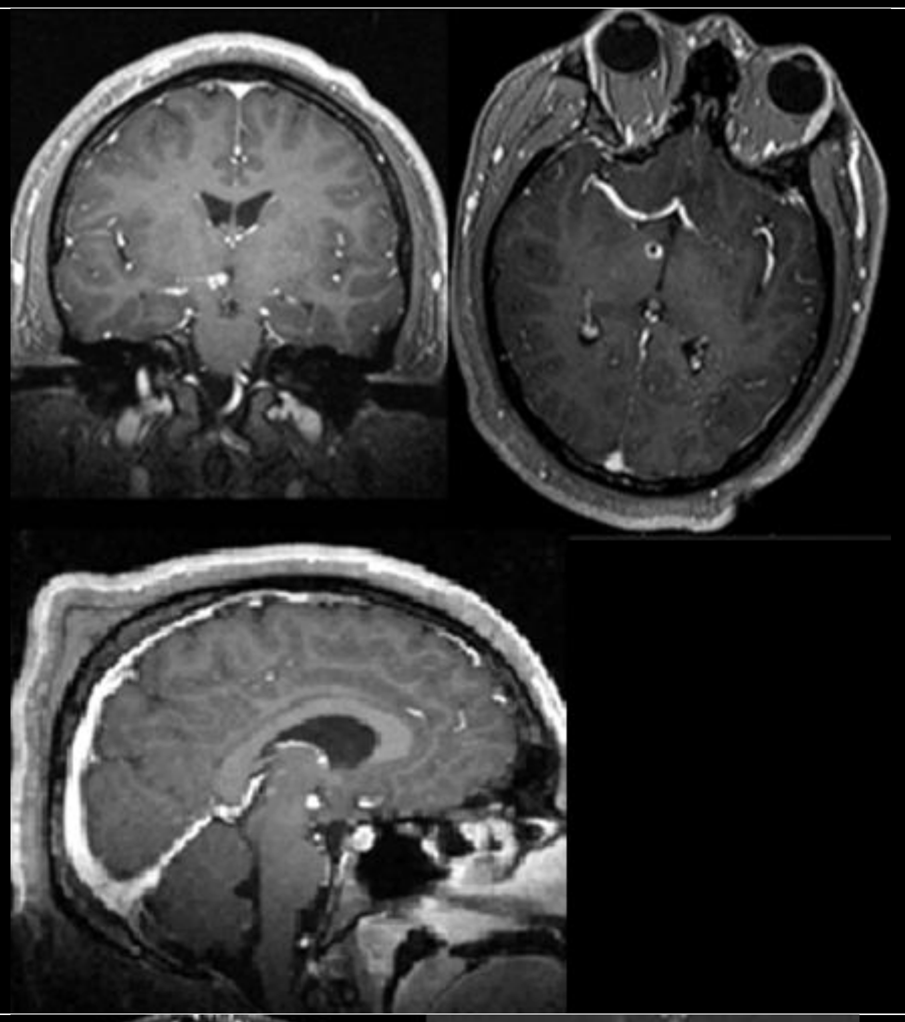 | 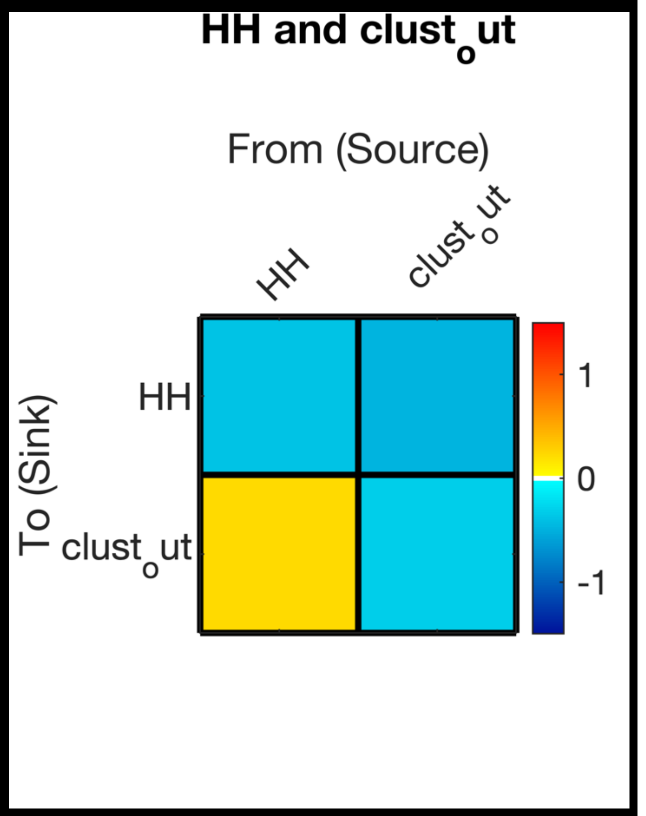  |
| 3 | 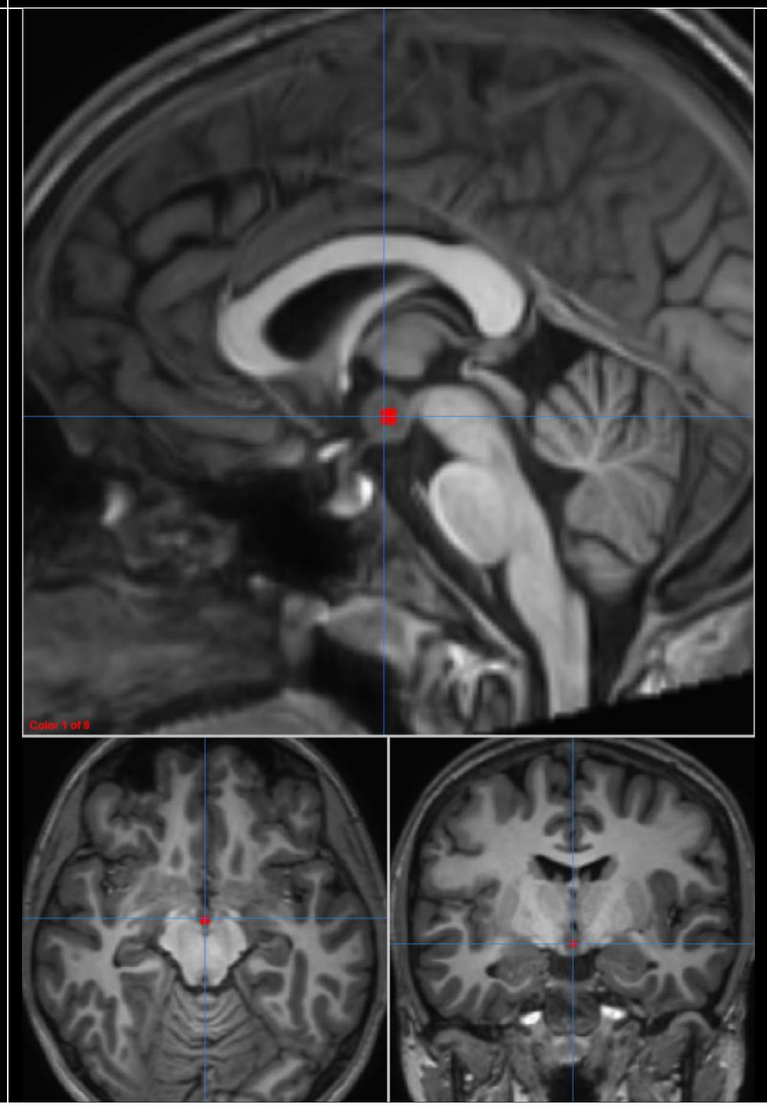 | 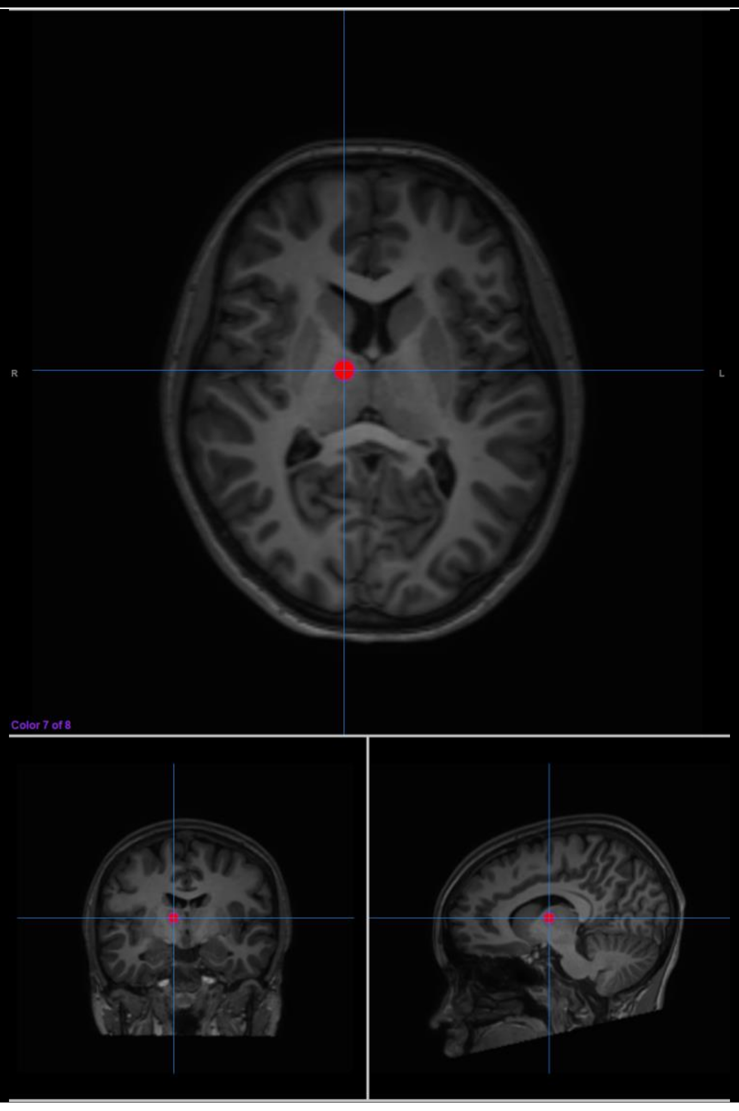 | 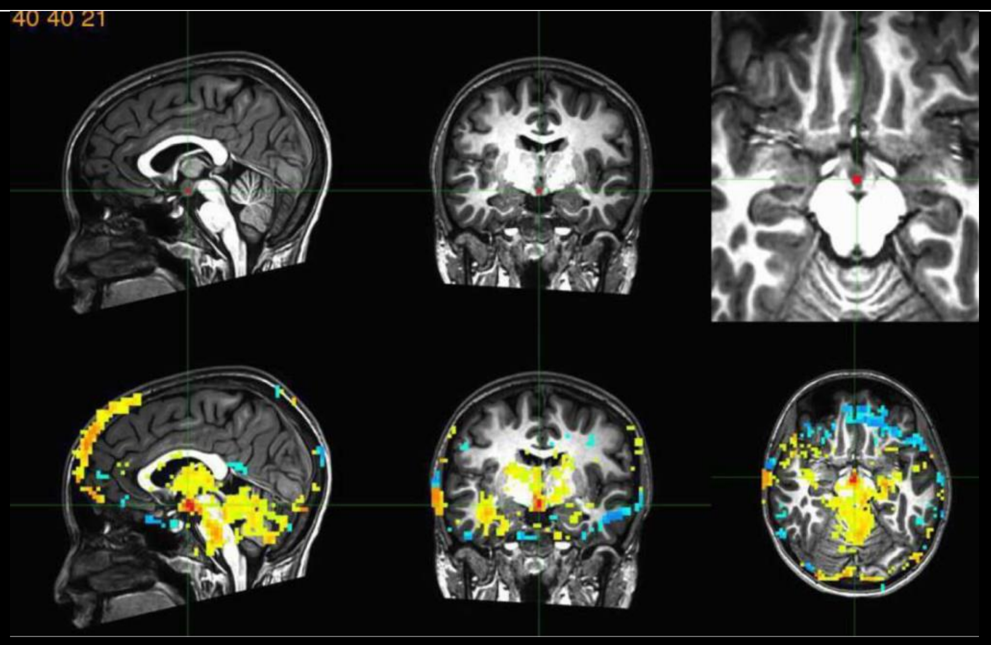  | 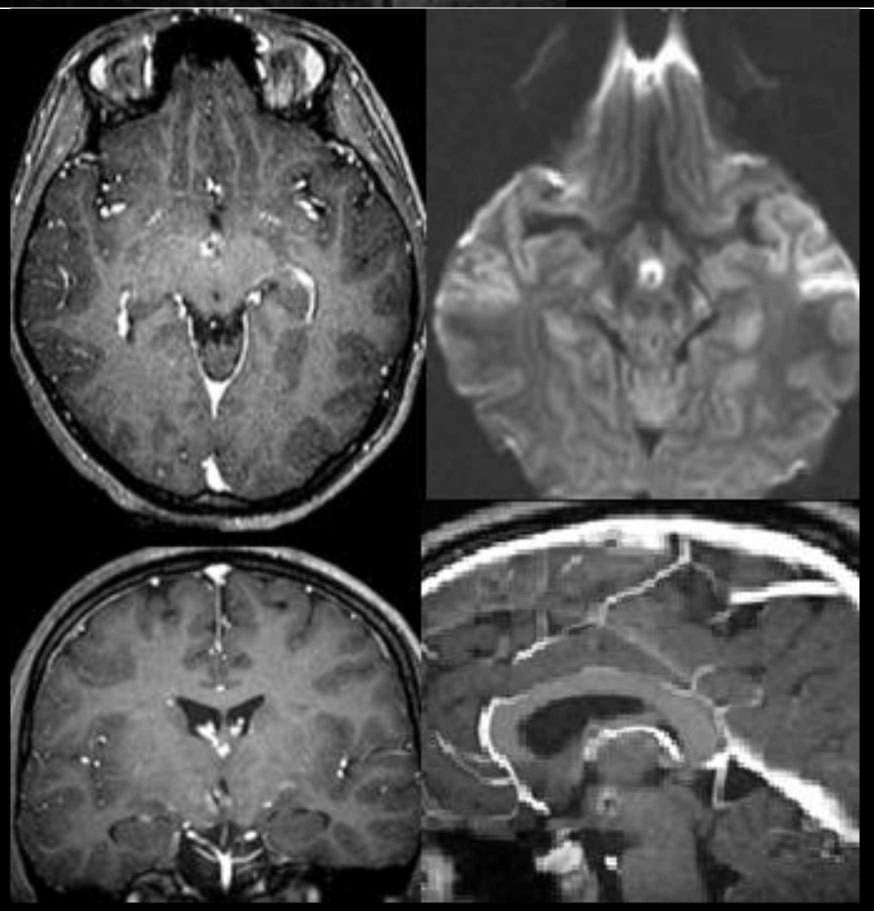 | 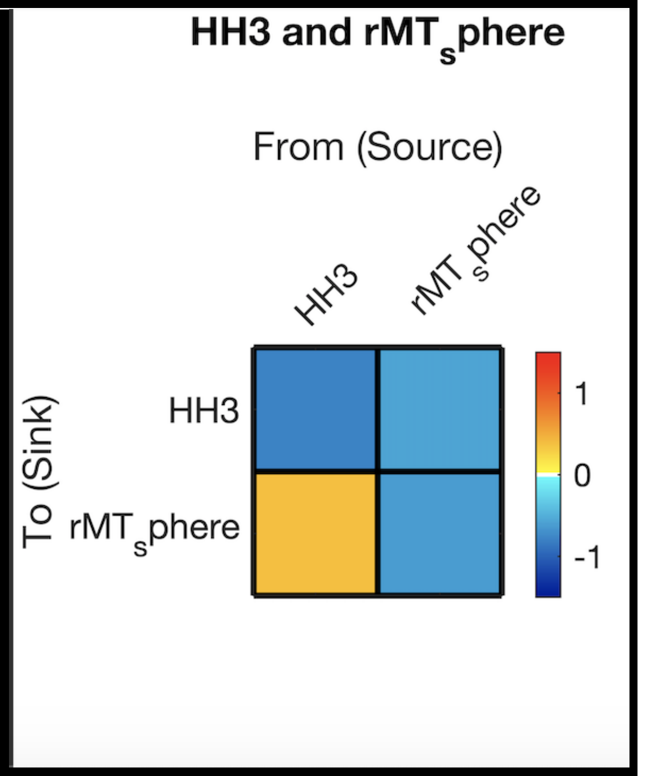  |

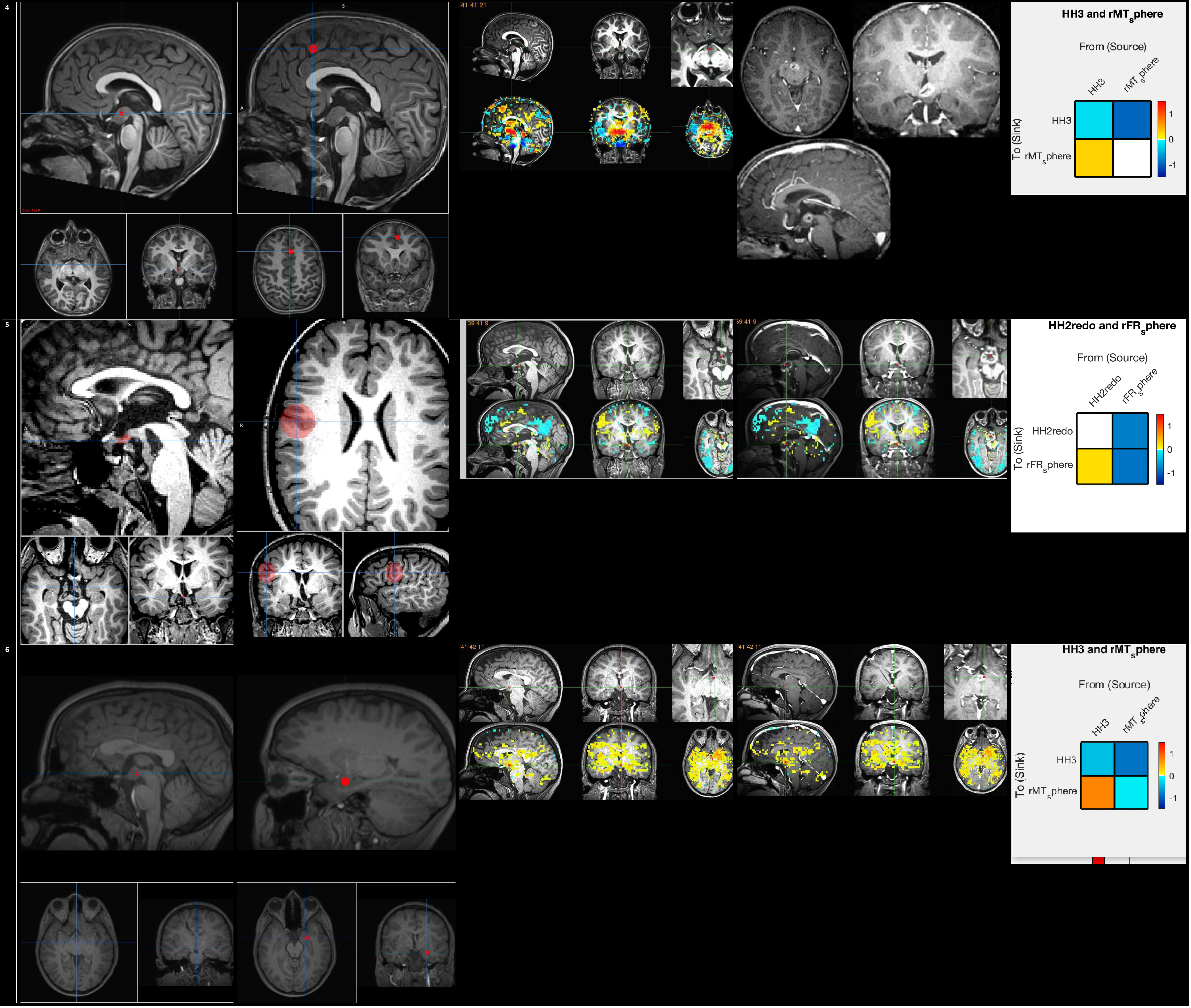
4
41 41 21
HH3 and rMT_sphere
From (Source)
HH3
rMT_sphere
To (Sink)
HH3
rMT_sphere
1
0
-1
5
39 41 9
39 41 9
HH2redo and rFR_sphere
From (Source)
HH2redo
rFR_sphere
To (Sink)
HH2redo
rFR_sphere
1
0
-1
6
41 42 11
41 42 11
HH3 and rMT_sphere
From (Source)
HH3
rMT_sphere
To (Sink)
HH3
rMT_sphere
1
0
-1

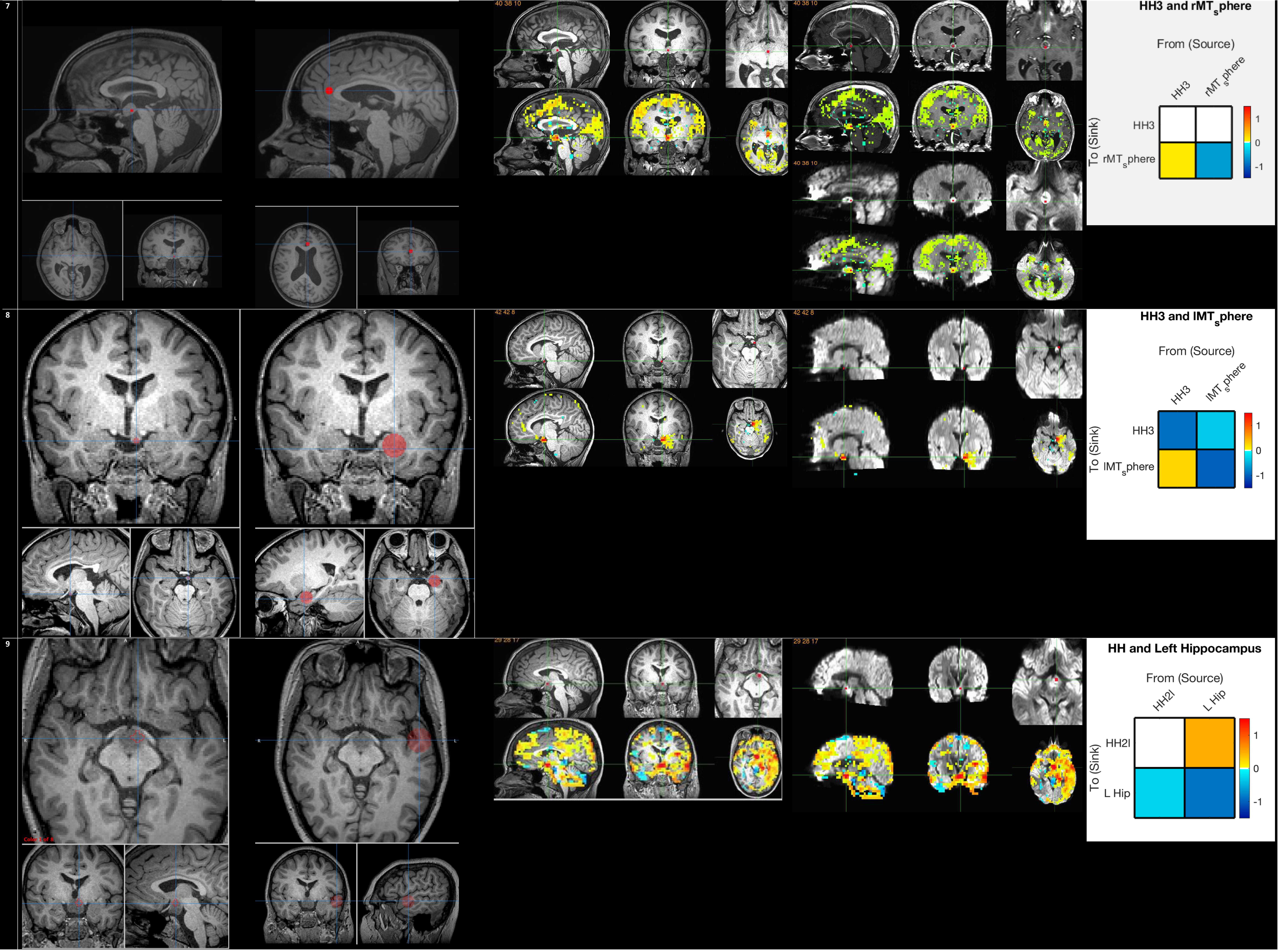
7
40 38 10
40 38 10
40 38 10
HH3 and rMT_sphere
From (Source)
HH3
rMT_sphere
To (Sink)
HH3
rMT_sphere
1
0
-1
8
42 42 8
42 42 8
HH3 and lMT_sphere
From (Source)
HH3
lMT_sphere
To (Sink)
HH3
lMT_sphere
1
0
-1
9
29 28 17
29 28 17
HH and Left Hippocampus
From (Source)
HH2l
L Hip
To (Sink)
HH2l
L Hip
1
0
-1

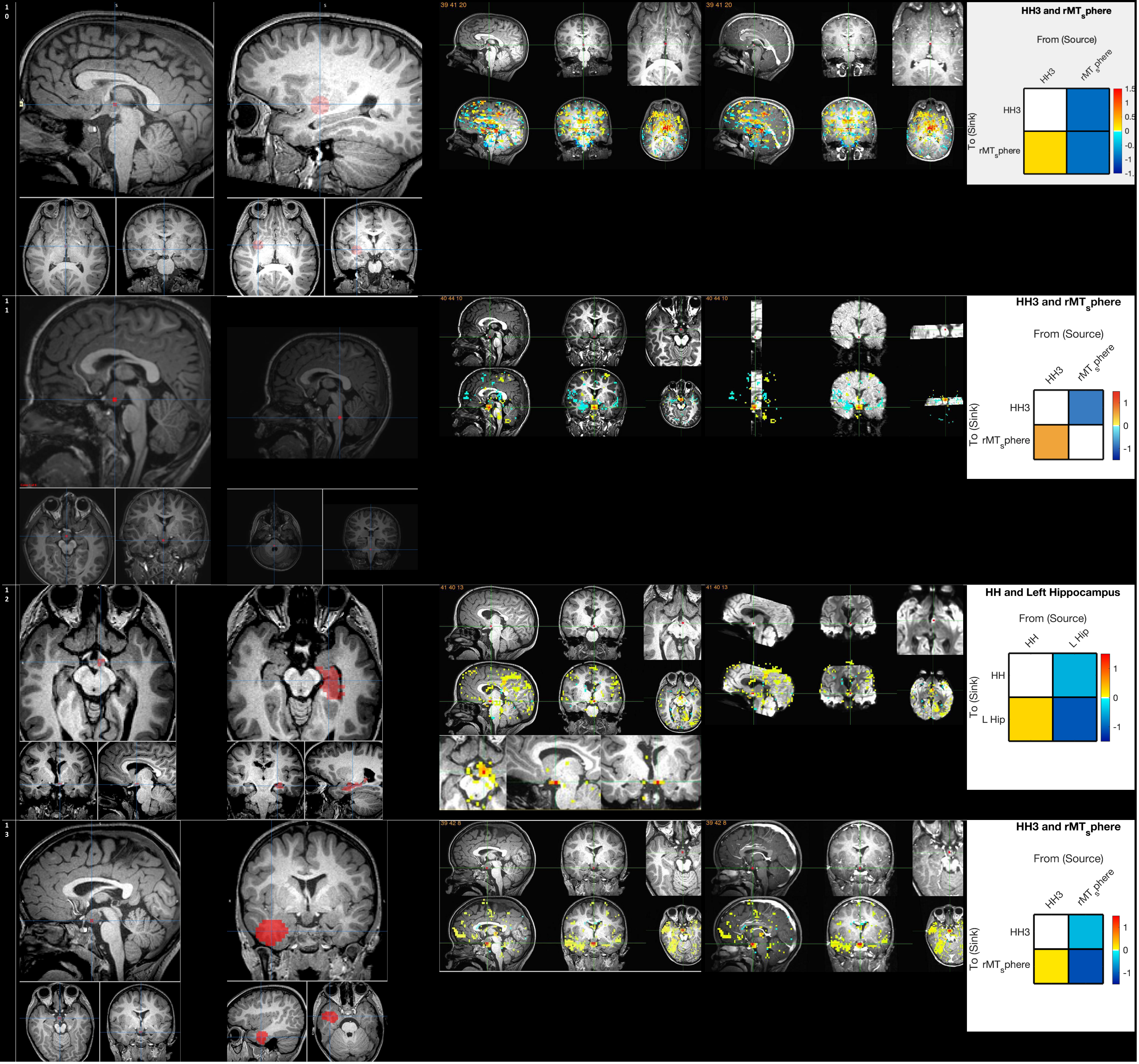

10
39 41 20
39 41 20
HH3 and rMT_sphere
From (Source)
HH3
rMT_sphere
To (Sink)
11
40 44 10
40 44 10
HH3 and rMT_sphere
From (Source)
HH3
rMT_sphere
To (Sink)
12
41 40 13
41 40 13
HH and Left Hippocampus
From (Source)
HH
L Hip
To (Sink)
13
39 42 8
39 42 8
HH3 and rMT_sphere
From (Source)
HH3
rMT_sphere
To (Sink)

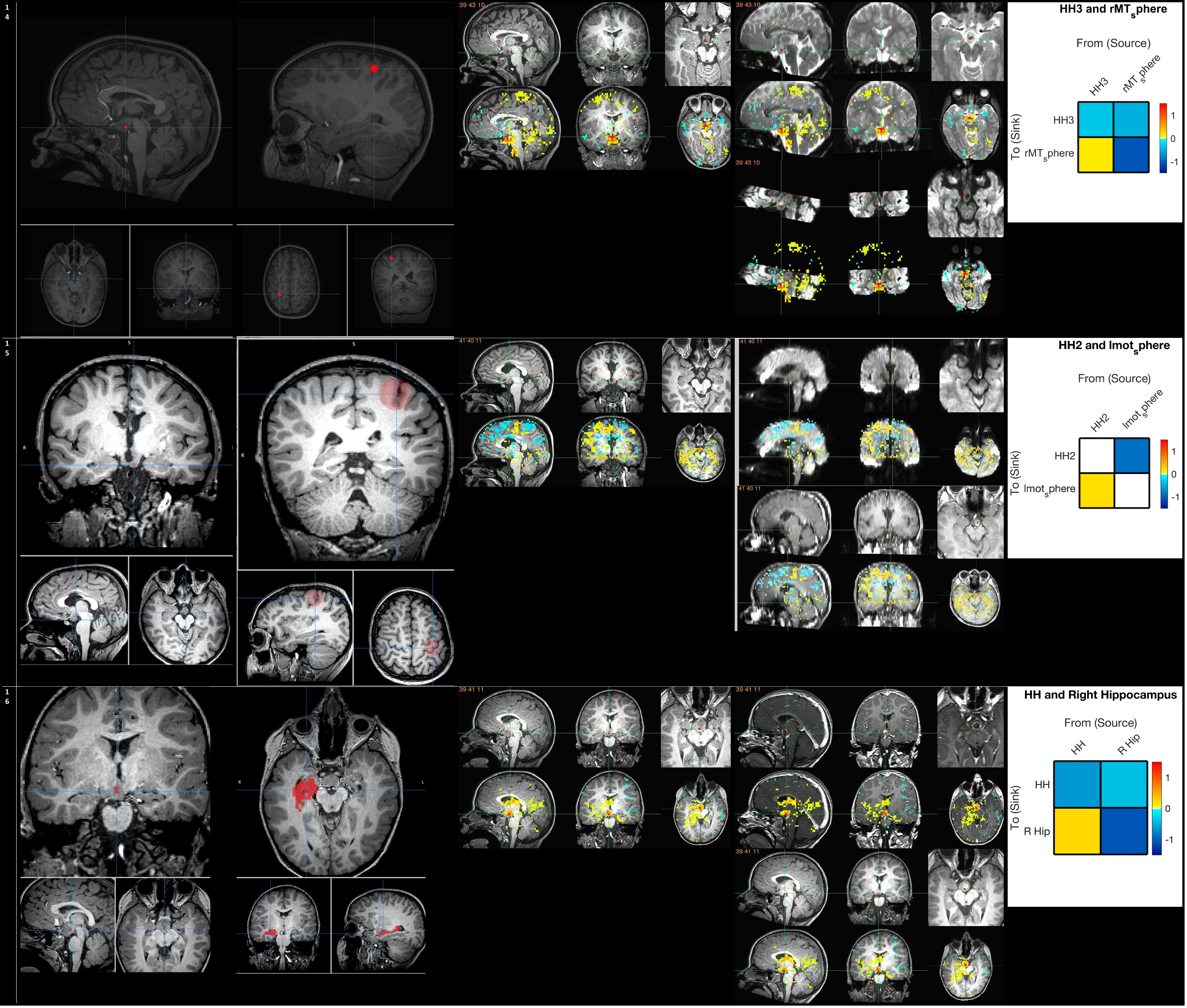

14
39 43 10
HH3 and rMTsphere
From (Source)
HH3
rMTsphere
To (Sink)
1
0
-1
15
41 40 11
HH2 and lmotsphere
From (Source)
HH2
lmotsphere
To (Sink)
1
0
-1
16
39 41 11
HH and Right Hippocampus
From (Source)
HH
R Hip
To (Sink)
1
0
-1

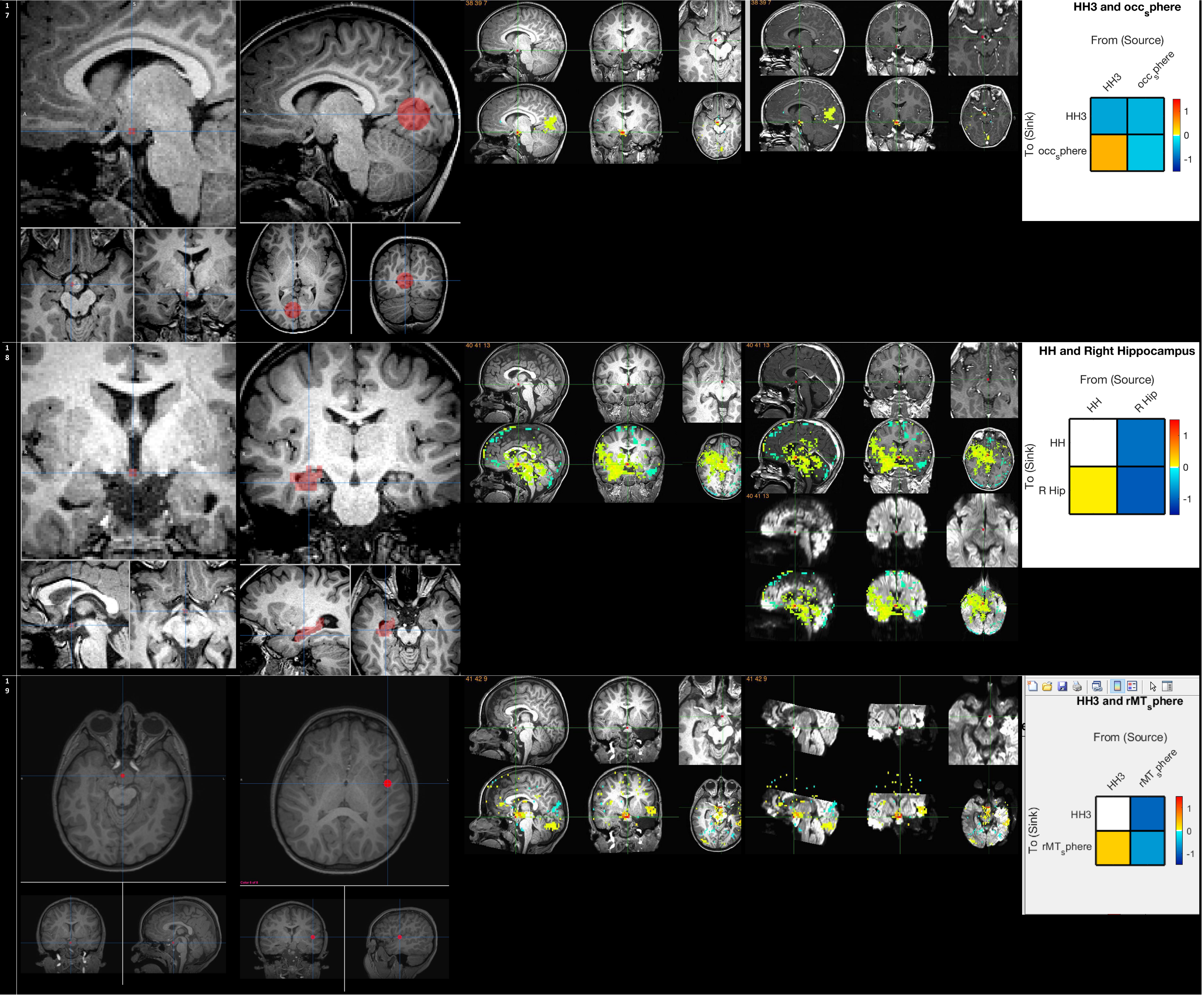

17
38 39 7
HH3 and occ_sphere
From (Source)
HH3
occ_sphere
To (Sink)
1
0
-1
18
40 41 13
HH and Right Hippocampus
From (Source)
HH
R Hip
To (Sink)
1
0
-1
19
41 42 9
HH3 and rMT_sphere
From (Source)
HH3
rMT_sphere
To (Sink)
1
0
-1

20

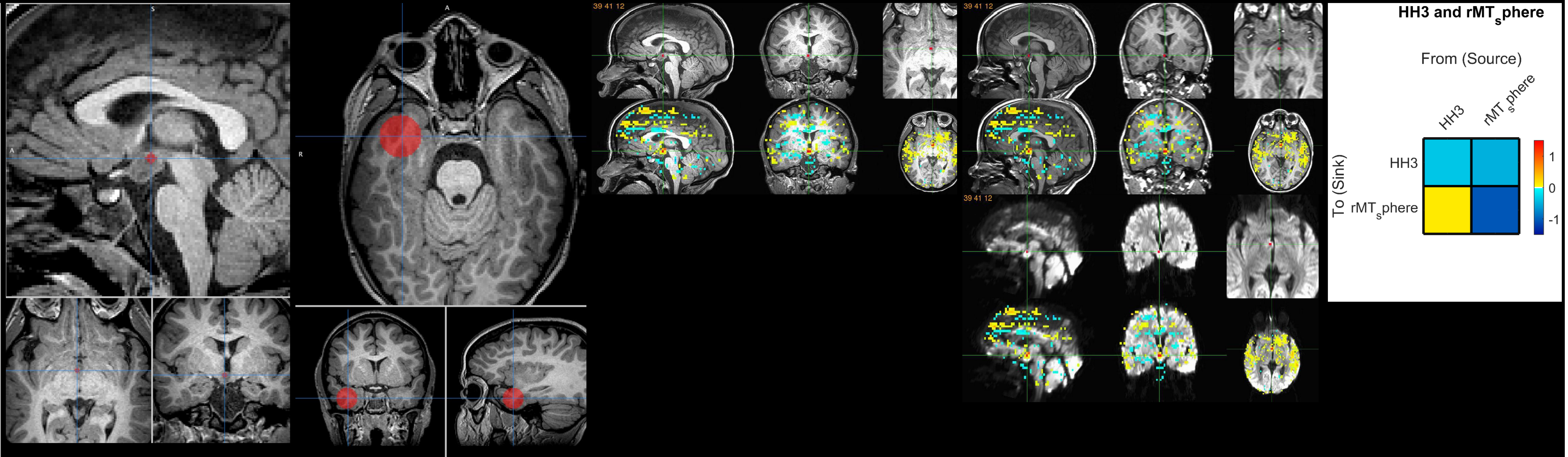
39 41 12
39 41 12
39 41 12
HH3 and rMT$_{s}$phere
From (Source)
HH3
rMT$_{s}$phere
To (Sink)
HH3
rMT$_{s}$phere
1
0
-1

21

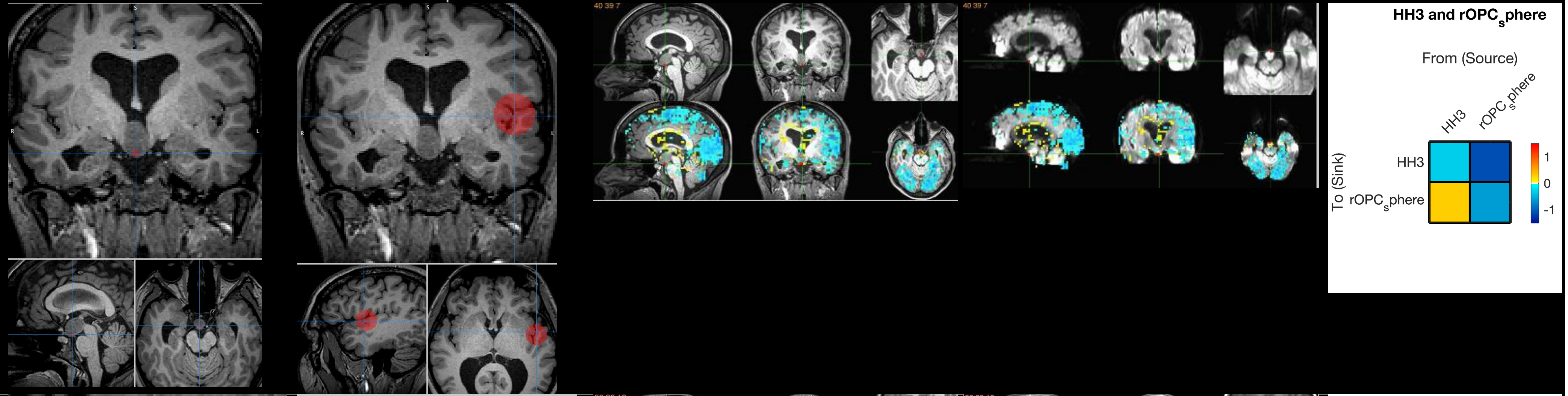
40 39 7
40 39 7
HH3 and rOPC$_{s}$phere
From (Source)
HH3
rOPC$_{s}$phere
To (Sink)
HH3
rOPC$_{s}$phere
1
0
-1

22

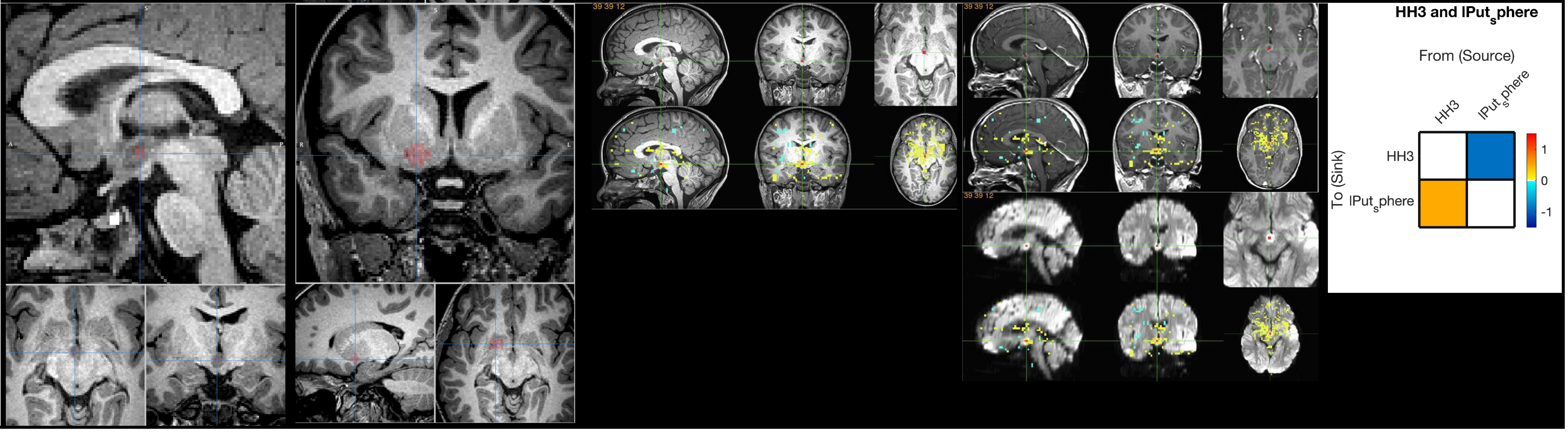
39 39 12
39 39 12
39 39 12
HH3 and lPut$_{s}$phere
From (Source)
HH3
lPut$_{s}$phere
To (Sink)
HH3
lPut$_{s}$phere
1
0
-1

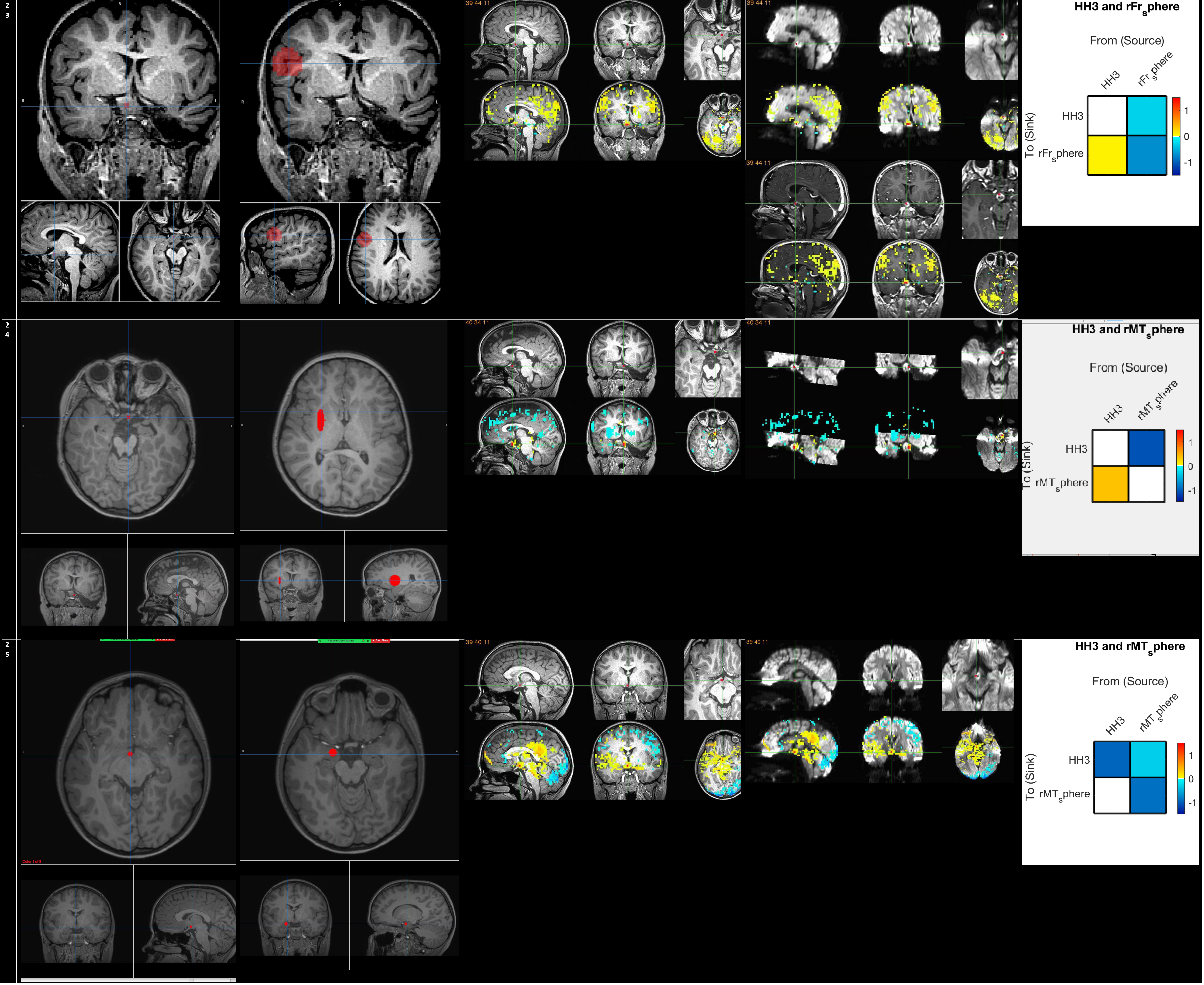

2
3
39 44 11
39 44 11
39 44 11
HH3 and rFr_sphere
From (Source)
HH3
rFr_sphere
To (Sink)
HH3
rFr_sphere
1
0
-1
2
4
40 34 11
40 34 11
HH3 and rMT_sphere
From (Source)
HH3
rMT_sphere
To (Sink)
HH3
rMT_sphere
1
0
-1
2
5
39 40 11
39 40 11
HH3 and rMT_sphere
From (Source)
HH3
rMT_sphere
To (Sink)
HH3
rMT_sphere
1
0
-1

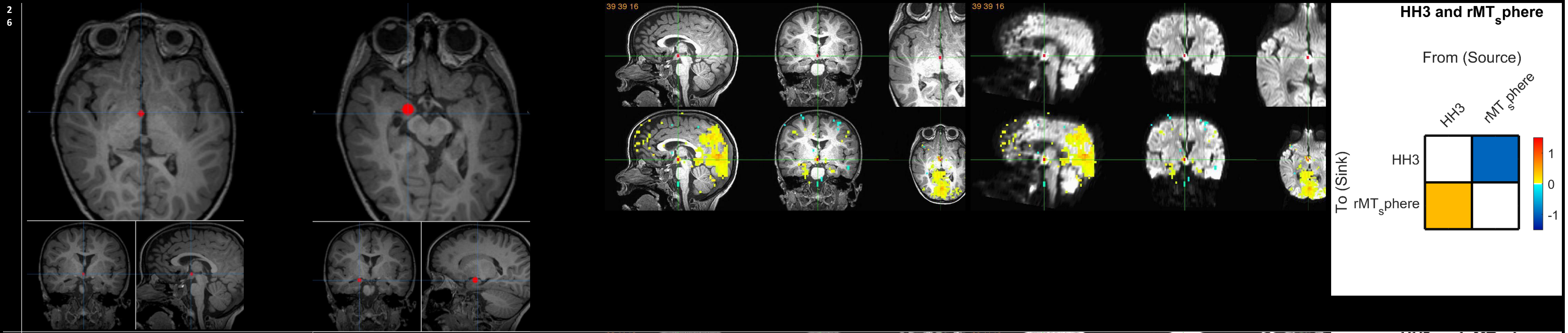
26
39 39 16
39 39 16
HH3 and rMT_sphere
From (Source)
HH3
rMT_sphere
To (Sink)
HH3
rMT_sphere
1
0
-1

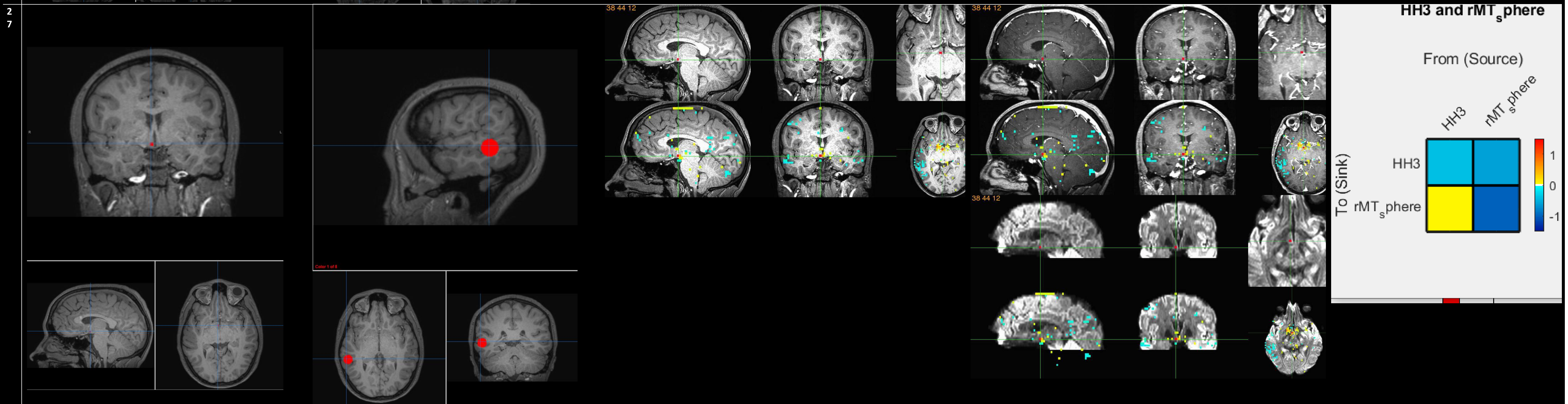
27
38 44 12
38 44 12
38 44 12
HH3 and rMT_sphere
From (Source)
HH3
rMT_sphere
To (Sink)
HH3
rMT_sphere
1
0
-1

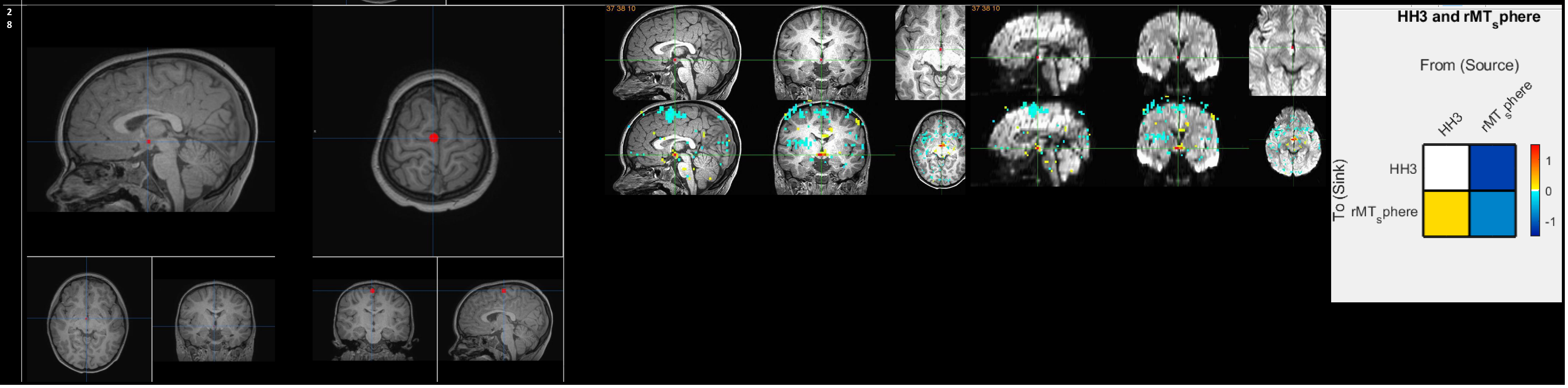
28
37 38 10
37 38 10
HH3 and rMT_sphere
From (Source)
HH3
rMT_sphere
To (Sink)
HH3
rMT_sphere
1
0
-1
