## Supplemental Table 2 for "Noninvasive Epileptogenic Signal Direction Determination by Effective Connectivity of Resting State Functional MRI - Independent of EEG"

**Table S2. Demographics, Seizure Frequency, and Engel Outcomes**

| **Patient** | **Gender M=1, F=2** | **Age at scan (yr) quartile range** | **Handed-ness R=1, L=2, ND=3** | **Pre-op seizure per month** | **Post-op seizure per month** | **Seizure improv count** | **Seizure improv %** | **Engel classification** |
| --- | --- | --- | --- | --- | --- | --- | --- | --- |
| 1 | 1 | (5.45-9.5) | 1 | 180 | 0 | -180 | 100 | Ia |
| 2 | 1 | (9.6-18.2) | 1 | 40 | 0 | -40 | 100 | Ia |
| 3 | 2 | (9.6-18.2) | 1 | 150 | 0 | -150 | 100 | Ia |
| 4 | 1 | (2.1-3.94) | 1 | 450 | 0 | -450 | 100 | Ia |
| 5 | 1 | (5.45-9.5) | 1 | 4 | 0 | -4 | 100 | Ia |
| 6 | 2 | (5.45-9.5) | 1 | 46 | 2 | -44 | 96 | Ib |
| 7 | 1 | (9.6-18.2) | 1 | 2 | 0 | -2 | 100 | Ia |
| 8 | 2 | (3.95-5.44) | 1 | 25 | 5 | -20 | 80 | Ib |
| 9 | 1 | (5.45-9.5) | 2 | 39 | 11 | -28 | 73 | Ib |
| 10 | 1 | (5.45-9.5) | 2 | 11 | 1 | -10 | 91 | Ib |
| 11 | 1 | (2.1-3.94) | 2 | 10 | 1 | -9 | 90 | Ib |
| 12 | 2 | (5.45-9.5) | 2 | 60 | 0 | -60 | 100 | Ia |
| 13 | 2 | (2.1-3.94) | 1 | 14 | 0 | -14 | 100 | Ia |
| 14 | 2 | (2.1-3.94) | 1 | 53 | 14 | -39 | 73 | Ib |
| 15 | 2 | (9.6-18.2) | 1 | 18 | 0 | -18 | 100 | Ia |
| 16 | 1 | (3.95-5.44) | 3 | 420 | 63 | -357 | 85 | Ib |
| 17 | 1 | (5.45-9.5) | 3 | 53 | 32 | -21 | 40 | Ib |
| 18 | 1 | (3.95-5.44) | 1 | 18 | 0 | -18 | 100 | Ia |
| 19 | 1 | (3.95-5.44) | 3 | 32 | 0 | -32 | 100 | Ia |
| 20 | 1 | (2.1-3.94) | 2 | 123 | 0 | -123 | 100 | Ia |
| 21 | 2 | (9.6-18.2) | 3 | 60 | 21 | -39 | 65 | Ib |
| 22 | 1 | (2.1-3.94) | 1 | 53 | 0 | -53 | 100 | Ia |
| 23 | 2 | (3.95-5.44) | 3 | 82 | 21 | -61 | 74 | Ib |
| 24 | 2 | (5.45-9.5) | 3 | 21 | 0 | -21 | 100 | Ia |
| 25 | 1 | (9.6-18.2) | 2 | 53 | 0 | -53 | 100 | Ia |
| 26 | 1 | (2.1-3.94) | 3 | 15 | 0 | -15 | 100 | Ia |
| 27 | 1 | (9.6-18.2) | 1 | 420 | 0 | -420 | 100 | Ia |
| 28 | 1 | (3.95-5.44) | 2 | 140 | 0 | -140 | 100 | Ia |

Abbreviations: Improv: improvement; ND: not determined; yr: year.

Age ranges are in quartiles.
