## Supplementary material for "Noninvasive Epileptogenic Signal Direction Determination by Effective Connectivity of Resting State Functional MRI - Independent of EEG": Supplmental Tables 3-5

**Table S3. Mann-Whitney Comparisons by Engel Outcomes**

|  | **Engel Ia (n=18)** | |  | **Engel Ib (n=10)** | |  |  |  |
| --- | --- | --- | --- | --- | --- | --- | --- | --- |
| **Factor** | **Mdn** | **[IQR]** |  | **Mdn** | **[IQR]** |  | **W** | ***P*-value** |
| Pre-operative seizure frequency | 46.25 | [17.5-135.6] |  | 49 | [28.2-58.1] |  | 92 | 0.94 |
| SOZ size | 1 | [1-1.75] |  | 1 | [1-1] |  | 76.5 | 0.53 |
| pZ size | 15.5 | [8-163.3] |  | 111 | [63-211.8] |  | 70 | 0.35 |
| Age (scan) | 5.47 | [3.9-11.1] |  | 5.44 | [4.3-8.7] |  | 92 | 0.94 |

Mann-Whitney tests were used due to violations of assumptions of normality and the relatively small and unevenly sized groups. Bonferroni corrected α is 0.013. Abbreviations: Mdn: median; IQR: interquartile range; SOZ: seizure onset zone; pz - propagation zone.

**Table S4. Correlations Among Patient Variables**

| **Variables** |  |  | | **r** | | ***P*-value** |
| --- | --- | --- | --- | --- | --- | --- |
| Age (scan) | - | SOZ size |  | -0.063 |  | 0.75 |
| Age (scan) | - | pZ size |  | 0.177 |  | 0.37 |
| Age (scan) | - | Pre-operative seizure frequency |  | 0.081 |  | 0.68 |
| Age (scan) | - | Pre-post seizure improvement (%) |  | -0.02 |  | 0.92 |
| SOZ size | - | SOZ-pZ |  | -0.136 |  | 0.49 |
| SOZ size | - | pZ-SOZ |  | -0.073 |  | 0.71 |
| SOZ size | - | SOZ-self |  | -0.09 |  | 0.65 |
| SOZ size | - | pz-self |  | -0.143 |  | 0.47 |
| pZ size | - | SOZ-pZ |  | -0.164 |  | 0.41 |
| pZ size | - | pZ-SOZ |  | -0.029 |  | 0.88 |
| pZ size | - | SOZ-self |  | -0.004 |  | 0.98 |
| pZ size | - | pz-self |  | 0.3 |  | 0.12 |

SOZ-pZ, pZ-SOZ, SOZ-self, and pZ-self are the posterior parameter estimates (strength values) of A matrix connections from the PEB analysis. Bonferroni corrected α is 0.004. Abbreviations: SOZ: seizure onset zone; pZ - propagation zone.

**Table S5. Comparisons by Sex**

|  | **Female (n=10)** | |  | **Male (n=18)** | |  |  |  |
| --- | --- | --- | --- | --- | --- | --- | --- | --- |
| **Factor** | **Mdn** | **[IQR]** |  | **Mdn** | **[IQR]** | **df** | **t** | ***P*-value** |
| SOZ-self | 0.39 | [-0.05-0.48] |  | 0.16 | [-0.1-0.32] | 26 | 0.86 | 0.4 |
| SOZ-pZ | 0.69 | [0.41-0.79] |  | 0.41 | [0.31--0.75] | 26 | 0.78 | 0.44 |
| pZ-SOZ | -1.6 | [-2.47-1.51] |  | -2.2 | [-3.28--1.4] | 26 | 0.62 | 0.5 |
| pZ-self | 0.36 | [-0.6-1.1] |  | 0.11 | [-0.75-1.1] | 26 | -0.35 | 0.7 |
|  |  |  |  |  |  |  | **W** | ***P*-value** |
| Pre-post seizure improvement (%) | 97.8 | [75.7-100] |  | 100 | [100-100] |  | 69.5 | 0.26 |
| The first four comparisons are of the posterior parameter estimates (strength values) of A matrix connections from the PEB analysis. All contained independent samples comparisons used student t-tests except for Pre-post seizure improvement (%), for which sample violated assumptions of normality, and thus a Mann-Whitney test was used. Bonferroni corrected α is 0.010. Abbreviations: Mdn: median; IQR: interquartile range; df: degrees of freedom; SOZ: seizure onset zone; pz - propagation zone. | | | | | | | | |
