## Supplemental Table 6 for "Noninvasive Epileptogenic Signal Direction Determination by Effective Connectivity of Resting State Functional MRI - Independent of EEG"

**Table S6. Clinical yield comparison**

|  | Vaudano 2021 |  |  | Us |  |  |  |
| --- | --- | --- | --- | --- | --- | --- | --- |
|  | Sample | less | Clinical Yield | Sample | less | Clinical Yield |  |
| Original sample | 35 |  |  | 36 |  |  | Comparable |
| Excluded: Concordant |  | 3 |  |  | na |  | Null relevance - patients with concordant fMRI activation maps are less likely to have an ambiguous SOZ location, lowering the clinical need for an EC analysis in the first place, thus we did not interpret it as a limitation |
| No surgery/iEEG |  | 4 |  |  | na |  | Null relevance – by design, our study sample included cases by surgery while the Vaudauno 2021 initial sample did not have this restriction |
| Excluded: Data File corruption* | na |  |  |  | 4 |  | Null relevance - Data corruption due to multiple data transfers between separate systems. Data were usable at time of patient’s clinical report for ICA and SL and otherwise should have been presumably usable for Directionality |
| Adjusted original Sample | 28 |  | 28/28 | 30 |  | 32/32 | Comparable |
| Excluded: Technical/no IED |  | 12 | 16/28 |  | na | 32/32 | Note: Vaudano 2021 does not detail ratio of technical vs. No IED or whether technical issues were related to data collection or e.g. file corruption.  Overall, our advancement is lower attrition due to data (un)usability; Directionality does not require IED capture, nor has technical issues associated with EEG-fMRI. Greatest threats to data integrity are patient movement and over sedation. |
| Excluded: Data quality/collection | na |  | 16/28 |  | 4 | 28/32 |  |
| Excluded: Null EEG-fMRI |  | 4 | 12/28 |  | 0 | 28/32 | Advancement: not required to capture IED-related fMRI activation, therefor not limited. Additionally, all patients had at least one pZ via SL that was outside the HH. |
| Excluded: Discordant EEG-fMRI |  | 6 | 6/28 |  | na | 28/32 |  |
| Surgery invalidated |  | 2 | 4/28 |  | 2 | 26/32 |  |
| Total: Subjects with DCM |  |  | 10/28 |  | 28 | 28/32 |  |
| Total: Subjects with DCM and surgery and/or iEEG |  |  | 6/28 |  | 28 | 28/32 |  |
| Total Validated accurate DCM-SOZ |  |  | 4/28 |  | 26 | 26/32 | In Vaudano 2021, patients had good outcomes when their DCM-SOZ and clinically indicated SOZ were concordant AND that region was surgically targeted, the patients with poor outcomes had a discordant DCM-SOZ and clinically indicated SOZ. Of note, the sample from Vaudano 2021 was relatively heterogenous group with focal epilepsy compared to our sample, though the majority were MRI positive |

Overall, due to constraints inherent with event-related DCM guided by EEG-fMRI, including substantial attrition related to technical constraints, lack of IED capture, and null fMRI activation associated with IED capture, the surgically-validated yield of this method is lower in the sample from Vaudano 2021 than in our sample.
